# Impact of vaccination on new SARS-CoV-2 infections in the UK

**DOI:** 10.1101/2021.04.22.21255913

**Authors:** Emma Pritchard, Philippa C. Matthews, Nicole Stoesser, David W. Eyre, Owen Gethings, Karina-Doris Vihta, Joel Jones, Thomas House, Harper VanSteenHouse, Iain Bell, John I Bell, John N Newton, Jeremy Farrar, Ian Diamond, Emma Rourke, Ruth Studley, Derrick Crook, Tim Peto, A. Sarah Walker, Koen B. Pouwels

## Abstract

The effectiveness of COVID-19 vaccination in preventing new SARS-CoV-2 infections in the general community is still unclear. Here, we used the Office for National Statistics (ONS) COVID-19 Infection Survey, a large community-based survey of individuals living in randomly selected private households across the UK, to assess the effectiveness of BNT162b2 (Pfizer-BioNTech) and ChAdOx1 nCoV-19 (Oxford-AstraZeneca; ChAdOx1) vaccines against any new SARS-CoV-2 PCR-positive tests, split according to self-reported symptoms, cycle threshold value (<30 versus ≥30) as a surrogate for viral load, and gene positivity pattern (compatible with B.1.1.7 or not). Using 1,945,071 RT-PCR results from nose and throat swabs taken from 383,812 participants between 1 December 2020 and 8 May 2021, we found that vaccination with the ChAdOx1 or BNT162b2 vaccines already reduced SARS-CoV-2 infections ≥21 days after the first dose (61%, 95% CI 54 to 68% versus 66%, 95% CI 60 to 71%, respectively) with greater reductions observed after a second dose (79%, 95% CI 65 to 88% versus 80%, 95% CI 73 to 85%, respectively). Largest reductions were observed for symptomatic infections and/or infections with a higher viral burden. Overall, COVID-19 vaccination reduced the number of new SARS-CoV-2 infections, with the largest benefit received after two vaccinations and against symptomatic and high viral burden infections, and with no evidence of difference between the BNT162b2 and ChAdOx1 vaccines.

## Introduction

On 8 December 2020, the UK was the first country to start a COVID-19 vaccination programme following the emergency use authorisation of the BNT162b2 mRNA vaccine (Pfizer-BioNtech) by UK’s Medicines & Healthcare Products Regulatory Agency^1^. Additional COVID-19 vaccines have since been approved, including the Oxford-AstraZeneca adenovirus-vector vaccine, ChAdOx1 nCOV-19 (termed here ChAdOx1)^2^, and more recently an mRNA-based COVID-19 vaccine developed by Moderna, mRNA-1273^3^. To date, most vaccinated individuals in the UK received one or two doses of the BNT162b2or ChAdOx1 vaccines, which are the vaccines focused on in the current study.

Initially, those in care homes, over 80 years old, and frontline health and social care workers were prioritised for vaccination^4^. Clinically vulnerable people and those ≥70 years were the next priority groups, followed by remaining adults in decreasing age order. As of 14 April, over 32 million (62%) UK adults (≥18 years old) had received at least one COVID-19 vaccine dose^5^, and mostly one dose only following the extension of the dosing interval to 12 weeks to maximise initial coverage^6^. UK inhabitants were invited to receive a COVID-19 vaccine independent of antibody status, although those testing PCR positive just before their scheduled vaccination had to reschedule their appointment to a later date to minimise the chances of an outbreak at vaccination sites.

Large randomised trials estimated efficacy against symptomatic laboratory-confirmed COVID-19 infection of 70% (95% CI 55% to 81%) after two ChAdOx1 doses^7^, and 95% (95% CI 90% to 98%) after two BNT162b2 doses^8^. Whilst trials provide unbiased effect estimates, trial participants may differ from the general population in many ways, and so it is essential to assess effectiveness in the community, particularly given differences between real-world vaccine deployment and the licenced dosing schedule. Comparing vaccine effectiveness in the community is also important as the trials used different outcome definitions (e.g. start of at-risk period 14^7^ vs 7^8^ days after the second dose) and populations (e.g. smaller proportion >55 years in the ChAdOx1 vaccine trial (12%^7^ vs 42% for BNT162b2^8^)).

Furthermore, both trials were largely conducted before the SARS-CoV-2 variant, B.1.1.7, became dominant in the UK^9^. This variant is more transmissible and potentially causes more severe disease^10-12^. Concerns have been raised that some of its defining mutations may affect the efficacy of vaccines and natural infection-derived immunity to (re)infection^13^. A subset of 8,534 participants from the initial ChAdOx1 trial were followed for a longer period to assess protection against different viral variants, but wide confidence intervals meant it was difficult to conclude whether efficacy was lower against B.1.1.7 (70%, 95%CI 44% to 85%) than other lineages (82%, 95%CI 70% to 89%)^14^.

Ongoing assessment of the effectiveness of different vaccines across different subgroups is critical – especially amongst older adults, who were underrepresented in the ChAdOx1 trials. Real-world studies are starting to appear, with an analysis from Israel estimating 92% (95%CI 88 to 95%) effectiveness against symptomatic PCR-confirmed infection ≥7 days after the second BNT162b2 dose^15^. A study among healthcare workers in England found an effectiveness of 70% (95% CI 55-85%) 21 days after a first dose and 85% (95% CI 74-96%) after a second dose of BNT162b2 against PCR-positive infections^16^. Another study assessing the early effectiveness of the BNT162b2 and ChAdOx1 vaccine in older adults (≥70 years) in England showed a single dose of either vaccine was ∼60% effective against symptomatic laboratory-confirmed infection and ∼80% effective against hospitalisation^17^. The evidence on effectiveness against asymptomatic infection is limited, with one study among 13,109 healthcare workers from Oxfordshire, UK, showing a 64% (95% CI 50 to 74%) reduction in any SARS-CoV-2 PCR-positive result following a single BNT162b2 or ChAdOx1 dose^9^. Another study among 3,950 healthcare workers, first responders, and other essential and frontline workers from the US estimated 80% (95%CI 59 to 90%) and 90% (95%CI 68 to 97%) vaccine effectiveness 14 or more days after 1 or 2 doses of the BNT162b2 or mRNA-1273 vaccines respectively^18^. Most recently, a study in 10,412 residents of long-term care facilities showed 65% and 68% protection against SARS-CoV-2 PCR-positive results 28-42 days after vaccination with ChAdOx1 and BNT162b2 vaccines respectively^19^.

However, existing studies have either investigated defined sub-populations^9 18 19^ or have relied on results from symptomatic testing programmes^15 17^, potentially leading to bias from vaccination status influencing test-seeking behaviour of cases not requiring healthcare. Large community-based studies where testing is done in a systematic manner (independent of both vaccination status and symptoms) are lacking. We therefore used the Office for National Statistics (ONS) COVID-19 Infection Survey (CIS) – a large community-based survey of individuals aged 2 years and older living in randomly selected private households across the UK – to assess the effectiveness of BNT162b2 and ChAdOx1 vaccines – as implemented in the UK – against any SARS-CoV-2 PCR positive test performed in the survey^20^, where RT-PCR tests were done on a fixed schedule, irrespective of symptoms, vaccine status and prior infection. We assessed vaccine effectiveness based on overall RT-PCR positivity, and split according to self-reported symptoms, cycle threshold (Ct) value (<30 versus ≥30) as a surrogate for viral load, and gene positivity pattern (compatible with B.1.1.7 or not).

## Results

### Characteristics of visits and new PCR-positives included in analysis

From 1^st^ December 2020 to 8^th^ May 2021, 383,812 individuals from 216,953 households provided 1,945,071 RT-PCR results from nose and throat swabs in the COVID-19 Infection Survey (median [IQR] 5 [4 to 6]), of which 12,826 (0.8%) were the first positive in an infection episode and 1,932,245 (99.3%) were negative. The characteristics at each visit where these swabs were taken, and hence included in analyses, are shown in **Supplementary Table 1**. The median (IQR) age at included visits was 55 years (40 to 68), 6% occurred in those reporting non-white ethnicity, 4% reporting patient-facing health/social care work or working in a care home (high priority group for vaccination), and 27% reporting a long-term health condition (priority group for vaccination).

We classified each visit according to vaccination status and previous infection (**Supplementary Table 2**), classifying time from vaccination empirically based on modelling days since first vaccination as a continuous non-linear effect (**Extended Data Fig. 1**). The baseline group was visits occurring >21 days before vaccination in those with no evidence of previous infection (1,012,808 visits, 10,721 new PCR-positives). 21,442 visits (105 PCR-positives) occurred in unvaccinated participants with evidence of previous infection, a median 125 days (IQR 106 to 161) from first positive (study antibody, study swab, or external test in the national programme) to included visit. As <4% visits in each vaccinated group occurred in individuals with evidence of previous infection before vaccination (**Supplementary Table 2**), visits were classified based on vaccination history alone. As there was insufficient data to estimate effects of vaccination dependent on previous infection status, we therefore estimate the effectiveness of vaccination as implemented in the UK. 137,575 visits (95 PCR-positives) occurred a median 16 days (IQR 7 to 30) after a second dose; among the 99,267 individuals that received a second dose, the median days between the first and second dose was 73 (IQR 63 to 77).

**Figure 1:**
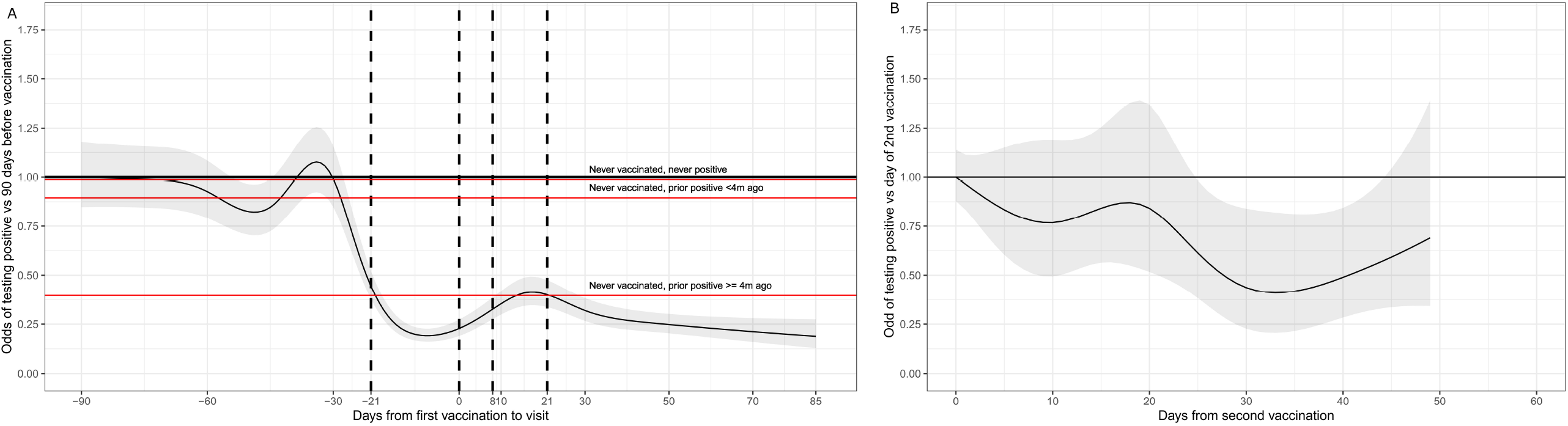
Distribution of Ct values (A) and percentage of symptoms (B) in new positive episodes by vaccination status. Number of visits with a positive test contributing to the plots by exposure group are 10,721 for ‘Not vaccinated, no prior positive, >21 days before vaccination’; 643 for ‘Not vaccinated, no prior positive, 1-21 days before vaccination’; 291 for ‘Vaccinated 0-7 days ago’; 441 for ‘Vaccinated 8-20 days ago; 530 for ‘≥21 days after 1^st^ dose, no second dose’; 95 for ‘Post second dose’; 76 for ‘Not vaccinated previously positive <4m ago’; 29 for ‘Not vaccinated, previously positive ≥4m ago. Boxplot inside violin shows the median, and upper and lower quartiles of the distribution, with whiskers extending from the hinge to the largest/smallest value no further than 1.5 times the inter-quartile range. Error bars in panel B represent 95% confidence intervals. Values given in **Supplementary Table 3**.

In new infections, Ct values (inversely related to viral load) increased with increasing time from first vaccination and number of doses (**Figure 1A; Supplementary Table 3**). The highest Ct values were in those who had received two vaccine doses, with a similar distribution to those not vaccinated but previously PCR or antibody-positive. Ct values were lowest in those not vaccinated and not previously PCR or antibody-positive.

The percentage of PCR-positive cases self-reporting symptoms was highest in those not vaccinated and not previously PCR or antibody-positive (58% >21 days prior vaccination), and lowest in those with two vaccine doses (17%) and those not vaccinated but previously PCR or antibody-positive ≥4 months ago (10%, **Figure 1B**). Well-recognised COVID-19 symptoms (cough, fever, loss of taste/smell) were most commonly reported in unvaccinated individuals and not previously PCR or antibody-positive.

### Impact of any COVID-19 vaccination on new infections

In unadjusted analyses, the percentage of positive PCR tests remained stable over the first 20 days following vaccination, but decreased from 21 days onwards regardless of having received one or two doses (**Extended Data Fig. 2**). Adjusting for multiple potential confounders, the vaccine effectiveness [(1-odds ratio)*100] against new PCR-positives, with or without symptoms, was 56% (95% CI 51 to 61%) in those 8 to 20 days after vaccination versus the baseline group, with no evidence of a difference versus those vaccinated 0 to 7 days ago (P=0.251). Vaccine effectiveness was 64% (95% CI 59 to 68%; P<0.001) in those ≥21 days since first vaccination with no second dose, marginally more than those vaccinated 8 to 20 days ago (P=0.066) (**Figure 2A, Supplementary Table 4;** coefficients for all factors in **Supplementary Table 5**). Odds of testing positive were reduced 72% (95% CI 70 to 75%) 1 to 21 days before first vaccination and 63% (95% CI 58 to 68%) 0 to 7 days post vaccination versus the baseline group.

**Figure 2:**
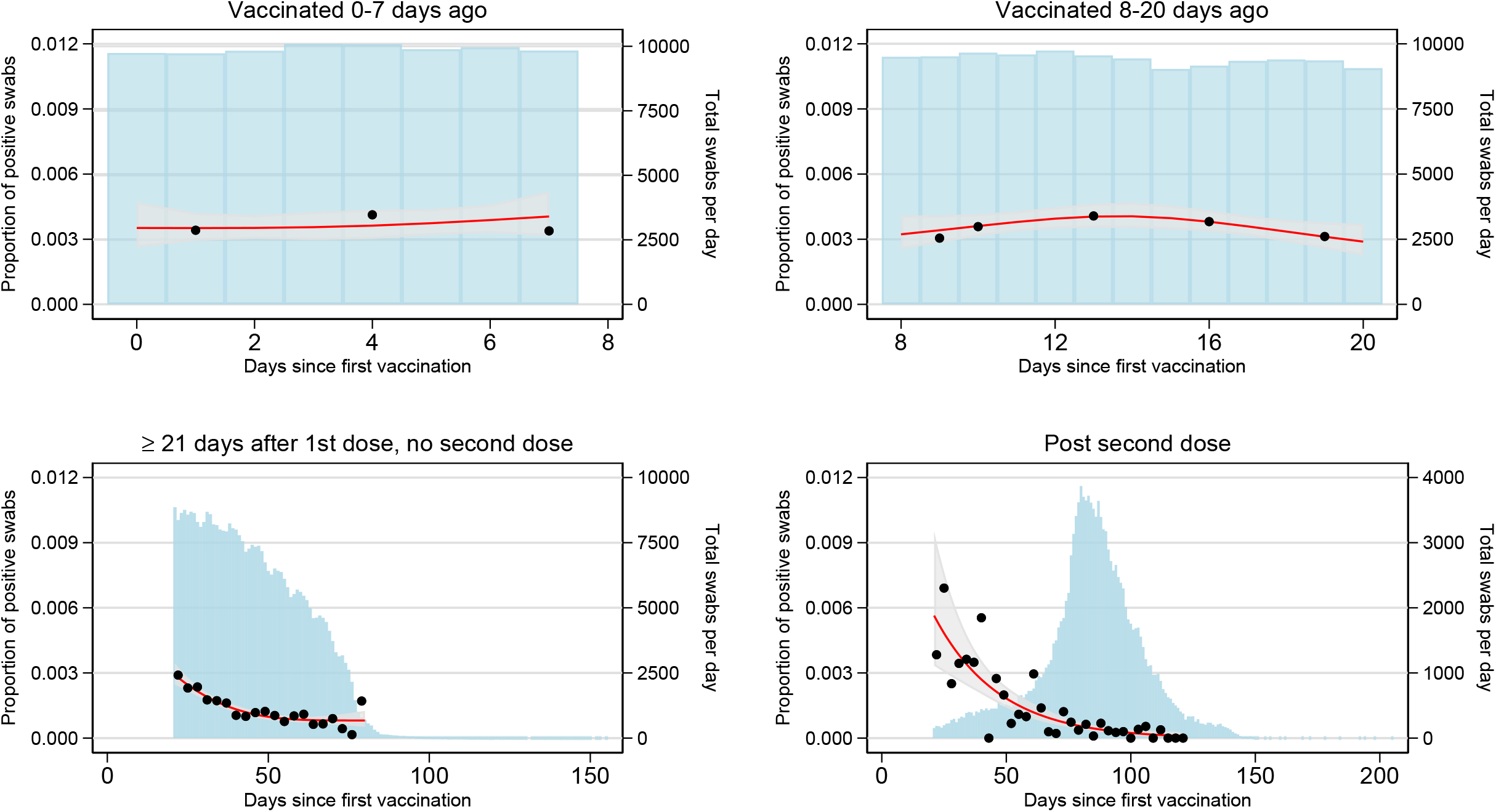
Adjusted odds ratios (95% CIs) for the effect of vaccination and prior positivity on: all positives (A), and positives split by Ct<30 or ≥30 (B), self-reported symptoms (C), and gene positivity pattern (D). All odds ratios were obtained from a generalised linear model with a logit link comparing to the reference category of “ Not vaccinated, not previously positive and ≥21 days before vaccination” and using clustered robust standard errors. *Not vaccinated, but with a positive antibody result in the study >90 days previously or a previous positive episode in the study Note: Odds ratios given in **Supplementary Table 4**. Number of visits underlying the models for the different outcomes are provided in **Supplementary Table 8**.

The largest vaccine effectiveness was estimated among those post second vaccine dose (80%, 95% CI 74 to 84%; P<0.001), significantly greater than for those having received only one dose ≥21 days previously (P<0.001). There was no evidence that reductions in odds of testing positive differed between having received two vaccine doses and not being vaccinated but PCR or antibody-positive >4m previously (P=0.523) (**Supplementary Table 4**).

The benefits associated with vaccination were much greater for infection episodes with Ct<30 as evidence of high levels of viral shedding compared with Ct≥30 (**Figure 2B**), with vaccine effectiveness against testing positive with Ct<30 estimated at 91% (95% CI 85 to 94%; P<0.001 post-second dose), a greater benefit compared to the 75% (69% to 79%) effectiveness following one dose ≥21 days ago (P<0.001) and with no evidence of difference versus those not vaccinated but PCR or antibody-positive >4m previously (P=1.00). Similarly, benefits associated with vaccination were much greater for self-reported symptomatic infection episodes (**Figure 2C**), with an estimated vaccine effectiveness against testing positive with self-reported symptoms of 95% (95% CI 90 to 97%; P<0.001) post-second dose, significantly higher than with one dose ≥21 days ago (P<0.001) (**Supplementary Table 4**), but again without evidence of difference versus those not vaccinated but PCR or antibody-positive >4m previously (P=1.00). In comparison, the estimated vaccine effectiveness against new infection episodes with no self-reported symptoms was 58% (95% CI 43 to 69%; P<0.001) post-second dose. Whilst overlapping, positives with Ct<30 also differed to positives reporting symptoms e.g. 4,731 (37%) of all positives had Ct <30 and symptoms reported, and 2,125 (17%) had Ct<30 and no symptoms reported (**Supplementary Table 3**). Effects of vaccination on infections compatible and not compatible with the B.1.1.7 variant appeared similar, but small numbers of positives in the latter group led to large uncertainty in estimates (**Figure 2D; Supplementary Table 4**).

### Impact of vaccination type on new infections

There was no evidence that vaccine effectiveness against new infections differed between the BNT162b2 and ChAdOx1 vaccine (**Figure 3A; Supplementary Table 6**) whether the vaccine was received 0 to 7 days ago (P=0.799), 8 to 20 days ago (P=1.00), ≥21 days ago (P=0.940), or post second dose (P=0.709). ≥21 days after the first dose, the effectiveness of BNT162b2 and ChAdOx1 vaccines was 66% (95% CI 60 to 71%) versus 61% (95% CI 54 to 68%), respectively and after 2 doses 80% (95% CI 73 to 85%) versus 79% (95% CI 65 to 88%) respectively. There was also no evidence that reductions in odds of new infections differed between those post second BNT162b2 dose and those not vaccinated but PCR or antibody-positive >4m previously (P=0.704). Effects were similar considering infections with Ct<30 and ≥30 (**Figure 3B**), and with and without self-reported symptoms (**Figure 3C**), with the impact of both vaccines attenuated to similarly degrees for infections with Ct≥30 and without self-reported symptoms.

**Figure 3:**
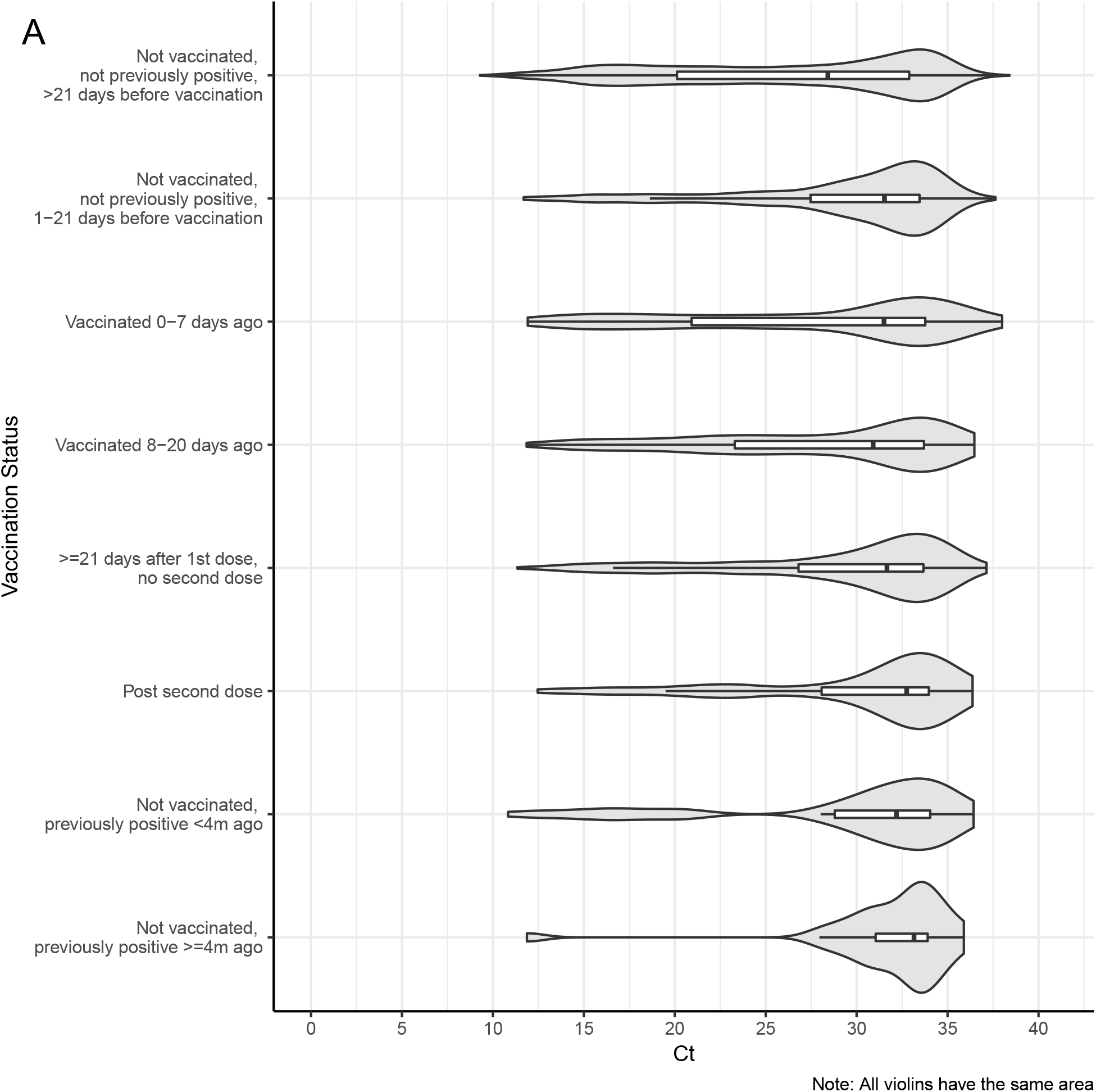
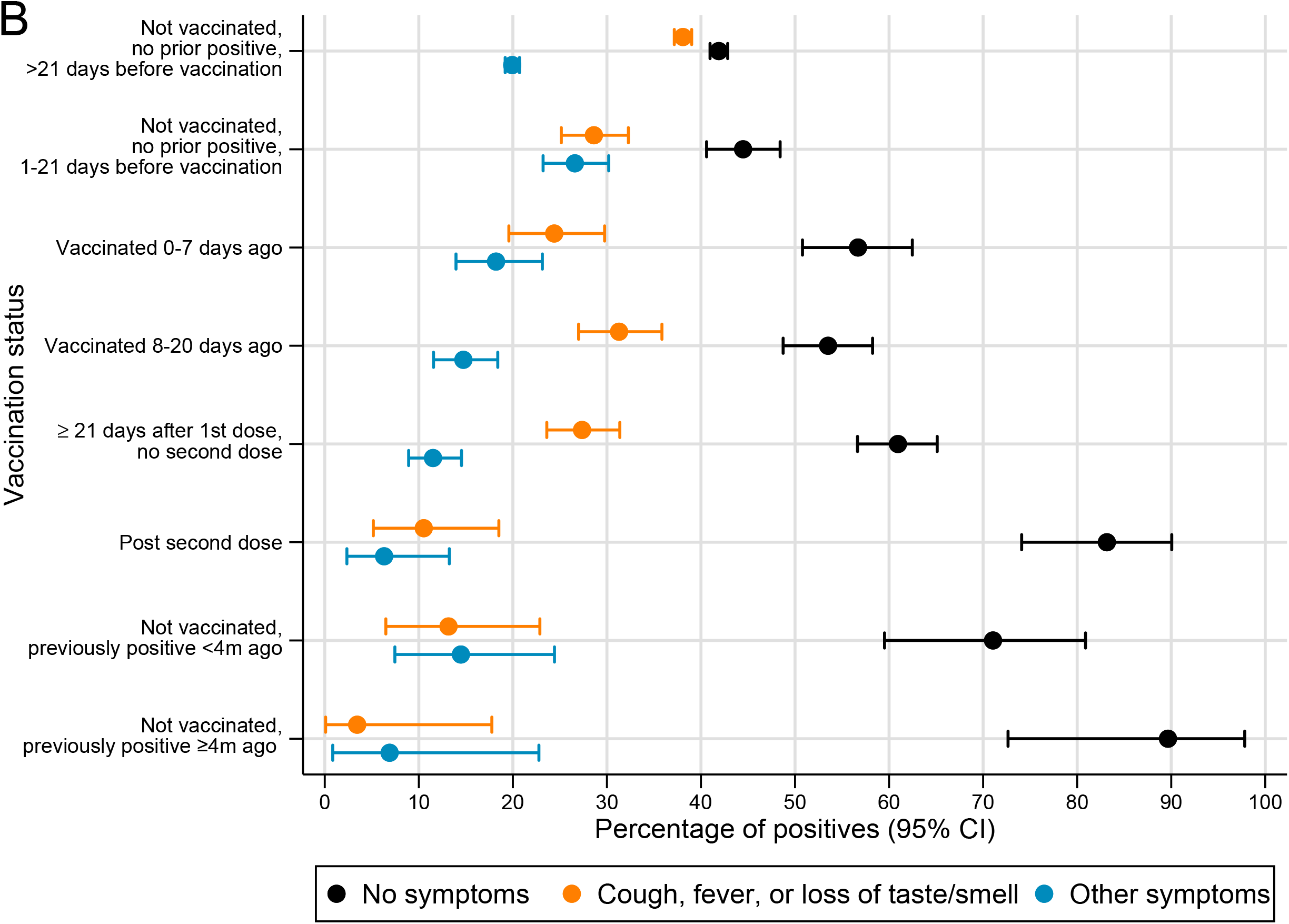
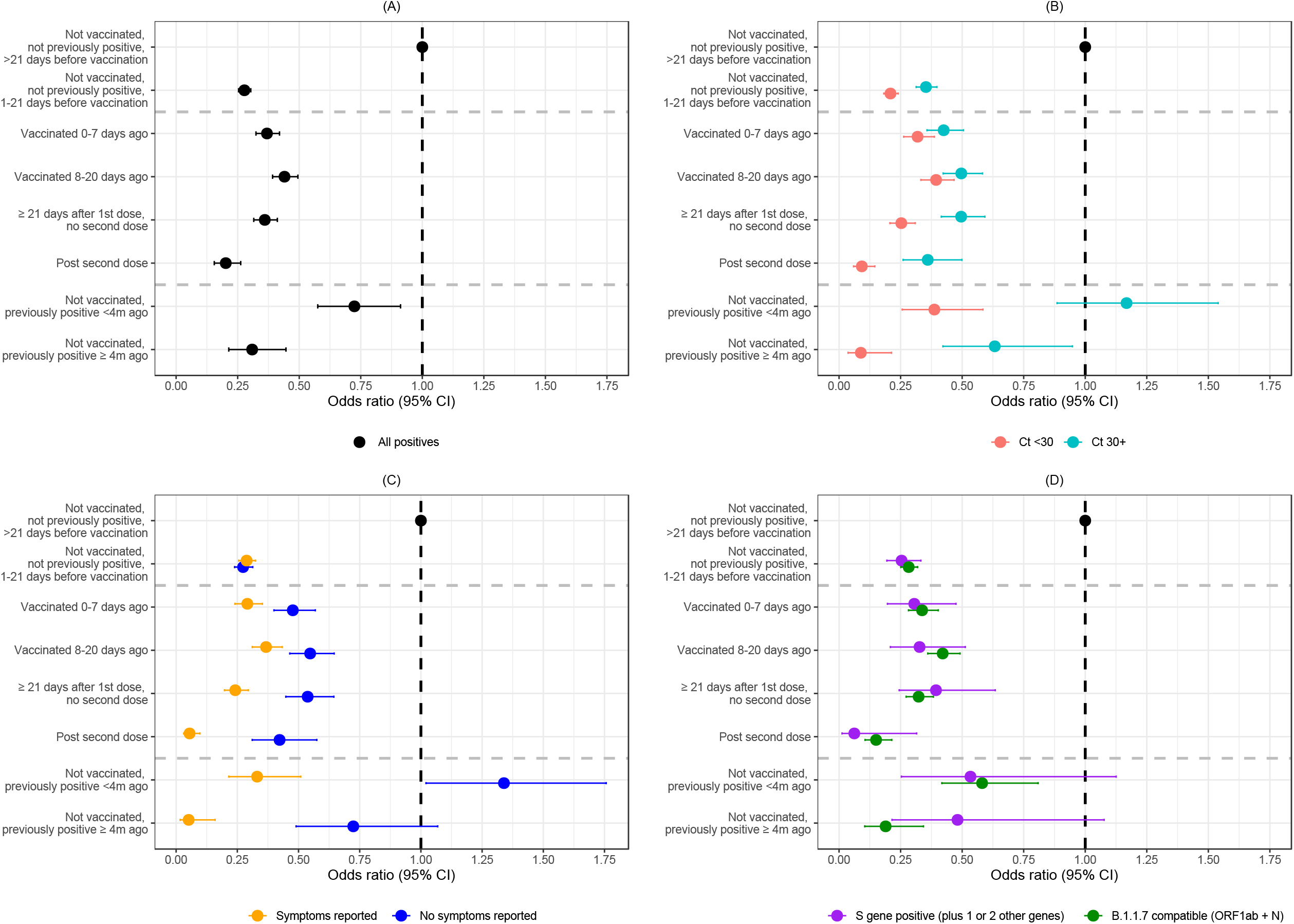
Adjusted odds ratios (95% CIs) for the effect of vaccination split by vaccine type and prior positivity on: all positives (A), and positives split by Ct<30 or ≥30 (B), self-reported symptoms (C). All odds ratios were obtained from a generalised linear model with a logit link comparing to the reference category of “ Not vaccinated, not previously positive and ≥21 days before vaccination” and using clustered robust standard errors. *Not vaccinated, but with a positive antibody result in the study >90 days previously or a previous positive episode in the study Note: Odds ratios given in **Supplementary Table 6**. Number of participants and visits underlying the models for the different outcomes are provided in **Supplementary Table 9**.

### Potential subgroup effects

There was evidence of differences in the effect of vaccination on new infection between those aged under or over 75 years (global heterogeneity for all vaccination terms P=0.011, **Figure 4A**). This was driven by greater benefits in those ≥21 days since first vaccination with no second dose, where reductions in odds were 72% in those aged ≥75 (95% CI 64 to 78% reduction) and 60% in those <75 (95% CI 54 to 65%) (interaction P=0.007). There was no evidence of differences in the effect of vaccination on new infection between those reporting or not reporting long-term health conditions (global heterogeneity for all vaccination terms P=0.897).

**Figure 4:**
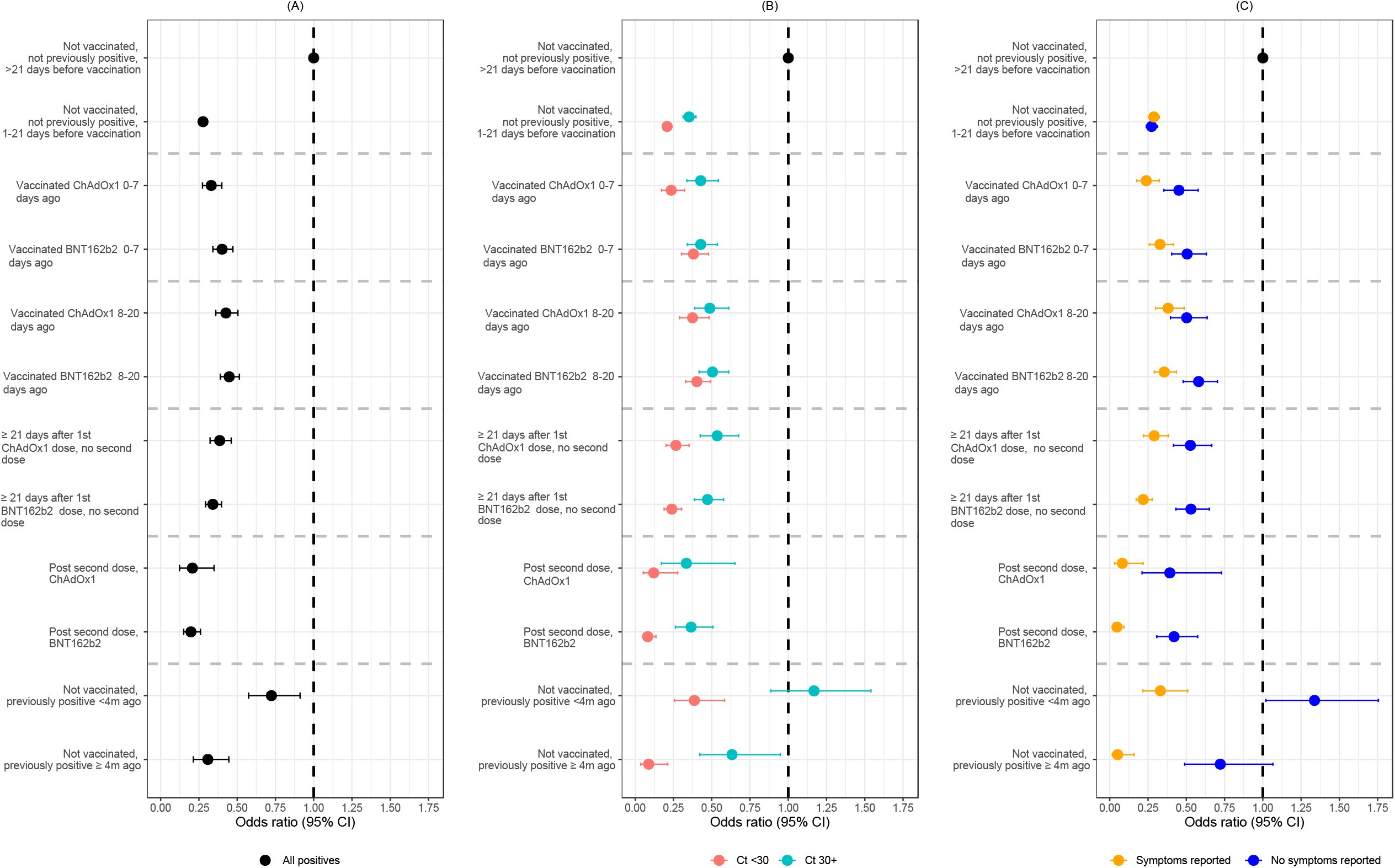
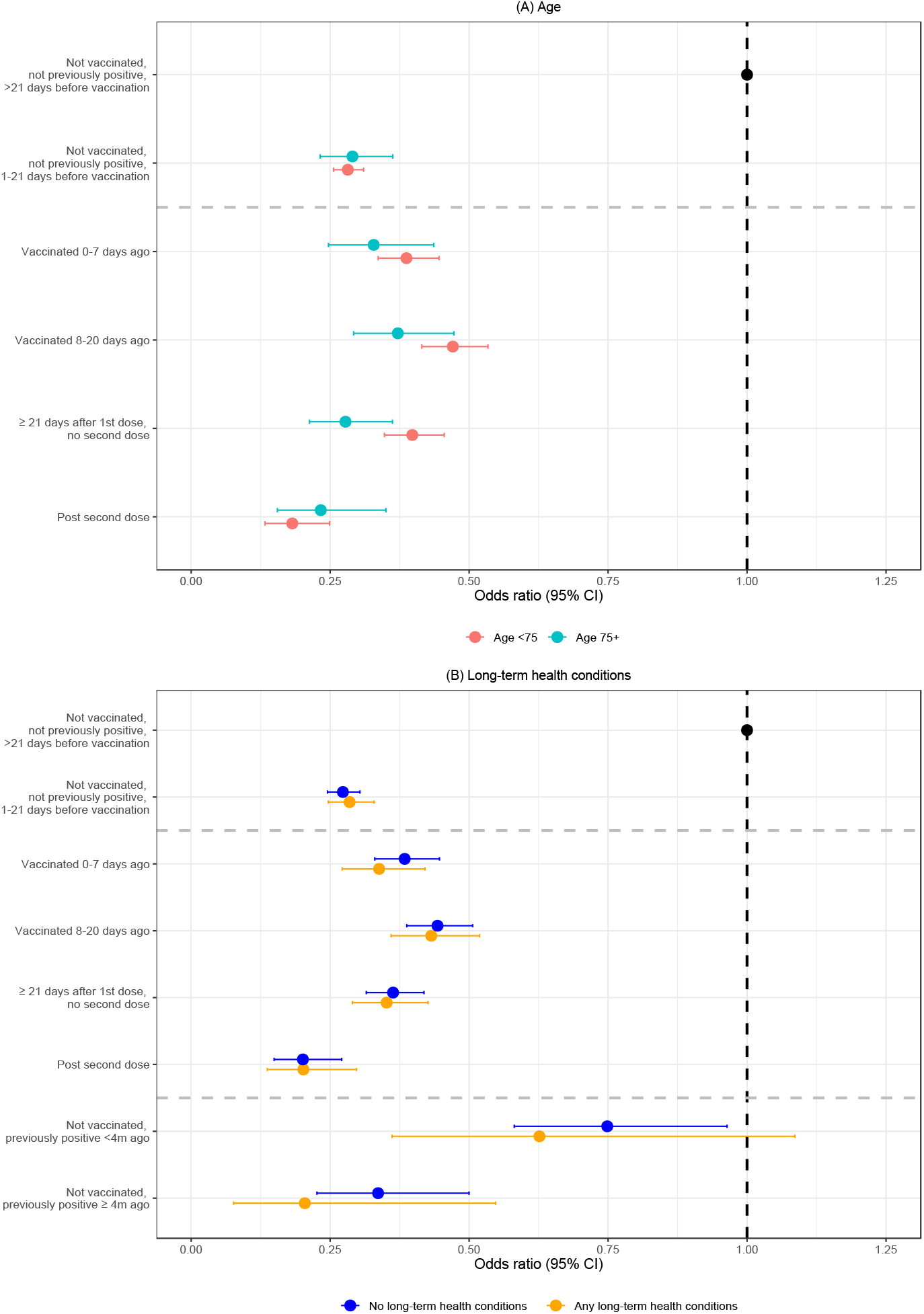
Adjusted odds ratios (95% CIs) for the effect of vaccination split by age <75 or 75+ (A) and long-term health conditions (B) on all positives. All odds ratios were obtained from a generalised linear model with logit link comparing to the reference category of “ Not vaccinated, not previously positive and ≥21 days before vaccination” and using clustered robust standard errors. *Not vaccinated, but with a positive antibody result in the study >90 days previously or a previous positive episode in the study. Number of participants and visits in the different potential subgroups are provided in **Supplementary Table 10 A** (by age) and **B** (by presence/absence of long-term health conditions). Note: Heterogeneity p-values (two-sided Wald test) for vaccination categories: Age p-value = 0.011, long-term health conditions p-value = 0.897. There were no positives in those aged ≥75 years in one previously infected exposure group, so these groups were excluded from subgroup analysis by age.

### Sensitivity analyses

While most potential confounders are unlikely affected by vaccination itself, work location, mode of travel to work, and contacts with care homes including visiting relatives, could theoretically be both confounders and affected through vaccination and prior infections through risk compensation. Sensitivity analyses excluding these factors resulted in nearly identical estimates for visits 1-21 days before vaccination, and very similar vaccine effectiveness ≥21 days since first vaccination with no second dose (64%, 95% CI 59 to 68% versus 60%, 95% CI 54 to 65%) and post second dose (80%, 95% CI 74 to 83% versus 77%, 95% CI 70 to 82%) (Supplementary Table 7).

The Ct threshold of <30> was selected on the basis of it being used in the UK for algorithms for review of low level positives at the laboratories where the PCR tests were performed and as a threshold for attempting whole genome sequencing. We also performed a sensitivity analysis with an arbitrary threshold of Ct <25> and found this increased the estimated vaccine effectiveness against ‘low Ct’ and ‘high Ct’ infections as lowering this threshold shifts both groups to a lower Ct value (higher viral load). For example, the estimated effectiveness ≥21 days after the first dose was 79% (95% CI 73 to 84%) for Ct <25 and 55% (95% CI 47 to 61%) for Ct≥25, while these values are 75% (95% CI 69 to 79%) and 50% (41 to 58%) with a threshold of 30, respectively.

## Discussion

The results from this large community surveillance study show that vaccination against COVID-19, with either ChAdOx1 or BNT162b2 -vaccine, significantly reduced the odds of individuals testing PCR-positive with a new SARS-CoV-2 infection, with greatest reductions in new infections observed in individuals with Ct<30 and self-reported symptoms, and in those who had received 2 vaccine doses. Reductions afforded by vaccination were similar to those not vaccinated but PCR or antibody-positive >4m previously. The protective effect of vaccination was more pronounced in infections with Ct<30 and with self-reported symptoms. There was no evidence of any difference in effectiveness between BNT162b2 and ChAdOx1 vaccines, or in those with long-term health conditions. We observed greater reductions in new infections in those aged ≥75 years versus those under 75 after one dose, but this difference was not apparent after 2 doses.

The main study strength is its design as a large-scale community survey recruiting from randomly selected private residential households, providing a representative sample of the UK general population. Participants are tested regardless of symptoms, allowing us to additionally consider vaccine effectiveness against infection without reported symptoms. The availability of Ct values allowed us to compare vaccine impact on viral loads, using Ct as a proxy^21^. Scheduled visits provide an unbiased sampling frame which we exploited for our logistic regression, rather than having to censor individuals at last tests in the study using time-to-event analyses, and assume all infections between visits were identified. Participants were asked about demographics, behaviours, and work, allowing us to control for a wide range of potential confounders that are unavailable in record linkage studies performed to date.^15^

The study design also has limitations, particularly with individuals tested initially at weekly and then monthly visits. Vaccination status was based on self-report in Northern Ireland, Wales and Scotland, potentially leading to some exposure misclassification. However, the vast majority of visits were for people living in England, where there was good agreement between self-reported and administrative vaccination data (98% on type and 95% on date). Antibody status was only measured in a subsample of study participants, meaning that some participants infected before joining the survey or not detected at survey visits will be misclassified as having no prior antibody-positive test. Similarly, any positive episodes occurring between visits and not captured by the national testing programme will be missed, leading to contamination of the “ not vaccinated, no previous PCR or antibody-positive” groups, possibly diluting the observed effects of vaccination. We used national testing programme positives only for exposure classification to avoid potential bias due to testing behaviour being influenced by vaccination and prior infection status. However, because participants can therefore only have a new positive outcome at scheduled visits, some of the “ new” positives episodes could have occurred sometime previously; we therefore stratified time from vaccination to reduce the impact of this. Older infections would be expected to have a higher Ct values, which might partly explain the differences between positives with Ct<30 and ≥30, at least shortly after vaccination.

Imperfect sensitivity of SARS-CoV-2 PCR tests may bias absolute risk, but would result in unbiased relative risk provided that outcome misclassification is non-differential to vaccination status and all non-cases are correctly classified (i.e. 100% specificity). PCR test specificity is likely very high (>99.99%)^12 20^, and therefore any bias here is expected to be small. Due to relatively small numbers of infections post-vaccination, power to detect differences between vaccine types and differential vaccine effectiveness in subgroups was relatively low.

An important potential issue with observational studies evaluating vaccine effectiveness is that individuals are not supposed to be vaccinated if they tested positive in the last 4 weeks,^22^ and individuals may reduce their number of contacts in response to the knowledge that they will soon receive a vaccination. We found that 643 individuals tested positive 1 to 21 days before receiving their vaccination – due to the design and logistics of the survey they might have received their test results after the date of vaccination – suggesting that ensuring social distancing at vaccination locations remains important. Rather than representing a biological effect, the reduced risk observed in the 21 days prior and 0-7 days after vaccination could be due to this reverse causality, specifically changes in behaviour due to either receiving the vaccination invitation letter, knowledge that individuals from their age or risk group are about to get vaccinated in their area or postponement of a planned vaccination visit due to a positive COVID-19 test in the 28 days prior to their scheduled vaccination appointment. In the hypothetical situation where all infections are detected immediately and adherence to guidance is perfect, there would be zero new infections observed 1 to 21 days before vaccination, emphasising the large temporal impact reverse causality can have. In theory, an unmeasured confounder could also explain the reduction in positive tests 1 to 21 days before receiving the vaccination compared to >21 days before receiving the vaccination; however, this would need to be a strong time-varying confounder that is at most weakly associated with calendar time as the latter was included in the model using a flexible tensor spline that modelled the interaction between non-linear effects of age and calendar time, and was also allowed to vary by region/country. Given this, it is much more likely that reverse causality underlies the lower odds of positive tests before and shortly after vaccination. Because a reduction in contacts and acquisition of infections in the week before vaccination will also reduce the likelihood of testing positive in the following week, it will be important for future studies trying to evaluate the effectiveness of vaccination to carefully construct the appropriate comparator. Here we used study visits from those that are not vaccinated, not previously positive, ≥21 days before vaccination as the baseline group to overcome these issues when estimating the impact of vaccination itself.

Our estimated effect of two vaccine doses on symptomatic infections is similar to that in the key Phase III clinical trials^7 8^, but slightly higher than other non-randomised studies which have considered this outcome but focused on specific populations or potentially affected by changes in test-seeking behaviour associated with vaccination^9 15 17 18,19^. Higher Ct in infections identified post vaccination has also been demonstrated in older adults in care homes^19^. Our estimated reduction in risk of new PCR-positives for those not vaccinated but infected >4m previously (69%) was slightly lower than the ∼80% (95% CI 75.4 to 84.5%) estimated elsewhere^23^, but we observed greater reductions against symptomatic (95%) and Ct<30 (91%) infections, suggesting this could be related to test seeking behaviour.

Consistent with two recent studies^9,14^, we found vaccination to be as effective against the B.1.1.7 variant as non-B.1.1.7 variants. Our study supports this in a broader population, including positives from individuals not reporting symptoms and for the BNT162b2 vaccine in addition to the ChAdOx1 vaccine. Our study had good power to estimate vaccine effectiveness against the B.1.1.7 variant as it was conducted over the period when B.1.1.7 became dominant in the UK. This is particularly relevant as the variant has now been detected in over 40 countries worldwide^24,25^, and the major Phase III vaccine trials were conducted before this strain was dominant^7 8^. We observed a slightly greater reduction in new infection episodes in those vaccinated and aged ≥75 years, compared with those <75 years – an effect that was no longer apparent after the second dose – potentially due to the combination of vaccination with reduced social contact in the former group. We currently do not have evidence of the vaccine being less effective in older individuals as seen elsewhere with natural re-infections^23^, although would note that, as described above, vaccine effectiveness also includes a non-biological behavioural component and there may be compensation for lower biological activity in older individuals with lower behavioural risk.

There was no evidence of any difference in effectiveness between BNT162b2 and ChAdOx1 vaccines post first or second doses. However, we cannot exclude the existence of small differences in vaccine effectiveness due to infrequent infections. There are very few direct head-to-head comparisons of both vaccines, especially after second doses, and it is important that differences between separate randomised trials are not directly attributed to the vaccine before considering other differences between trials, including different outcome definitions, populations, and circulating SARS-CoV-2 variants.^7,8^

Similar to other studies^8 9 16 18^, we found greater reductions in new positives after two vaccine doses compared with one dose, particularly in reducing infections with self-reported symptoms and low Ct/high viral load. In the UK, the interval between vaccine doses was extended to 12 weeks to maximise initial coverage and reduce hospitalisations and/or deaths; our findings highlight the importance for increased protection of individuals getting the second vaccine dose. Nonetheless, the significant reduction in positivity after only one dose supports the decision to maximise initial vaccination coverage.

While some infections, particularly those with Ct≥30, could represent historical infections contracted prior to vaccination given the timescales and prior negatives post vaccination, some will undoubtedly reflect new infections after vaccination. Together with other evidence, this suggests that vaccination does not completely prevent infection following virus exposure, yet minimises progression to more severe infection^15^. The fact that vaccinated individuals can still be infected, even if predominantly with lower viral burden and/or asymptomatic infections, means that onwards transmission remains a possibility, albeit at lower efficiency^26^. Overall, approximately one fifth of infections with Ct<30 were from individuals not reporting symptoms during their episode. These infections could be particularly relevant for transmission as those individuals may not be aware of their infection status despite having relatively high viral load. Maintaining measures such a social distancing may therefore still be needed to control virus spread until enough of the population is vaccinated.

We have also shown two vaccine doses to be as effective as prior natural infection. This could be an important consideration during policy development over COVID-status certification or so-called COVID passports, and supports considering both prior PCR or serological testing and vaccination data for this^27^.

Looking forward, one key question will be whether immunisation offers long-term protection against COVID-19. A recent study showed the rate of waning and longevity of neutralising antibodies varies greatly amongst individuals with prior COVID-19 infection and suggested that, if similar rates of waning are seen after vaccination or new variants that render vaccines less effective emerge and spread effectively (such as B.1.351 or P.1, which were too rare to assess in our study), more frequent vaccine administration is likely needed^28^. Overall, we have shown COVID-19 vaccination to be effective in reducing the number of new SARS-CoV2 infections, with the greatest benefit received after two vaccinations, and against symptomatic and high viral burden infections, and no difference in efftiveness between the BNT162b2 and ChAdOx1 vaccines.

## Data Availability

Data are still being collected for the COVID-19 Infection Survey. De-identified study data are available for access by accredited researchers in the ONS Secure Research Service (SRS) for accredited research purposes under part 5, chapter 5 of the Digital Economy Act 2017. For further information about accreditation, contact or visit the SRS website.

## Acknowledgements

This study is funded by the Department of Health and Social Care with in-kind support from the Welsh Government, the Department of Health on behalf of the Northern Ireland Government and the Scottish Government. EP, KBP, ASW, TEAP, NS, DE are supported by the National Institute for Health Research Health Protection Research Unit (NIHR HPRU) in Healthcare Associated Infections and Antimicrobial Resistance at the University of Oxford in partnership with Public Health England (PHE) (NIHR200915). ASW and TEAP are also supported by the NIHR Oxford Biomedical Research Centre. EP and KBP are also supported by the Huo Family Foundation. ASW is also supported by core support from the Medical Research Council UK to the MRC Clinical Trials Unit [MC_UU_12023/22] and is an NIHR Senior Investigator. PCM is funded by Wellcome (intermediate fellowship, grant ref 110110/Z/15/Z) and holds an NIHR Oxford BRC Senior Fellowship award. DWE is supported by a Robertson Fellowship and an NIHR Oxford BRC Senior Fellowship. The views expressed are those of the authors and not necessarily those of the National Health Service, NIHR, Department of Health, or PHE. The funder/sponsor did not have any role in the design and conduct of the study; collection, management, analysis, and interpretation of the data; preparation, review, or approval of the manuscript; and decision to submit the manuscript for publication. All authors had full access to all data analysis outputs (reports and tables) and take responsibility for their integrity and accuracy.

We are grateful for the support of all COVID-19 Infection Survey participants and the COVID-19 Infection Survey team.

## Author Contributions

The study was designed and planned by ASW, JF, JB, JN, IB, ID and KBP and is being conducted by ASW, IB, RS and ER. This specific analysis was designed by ASW, KBP, PCM, NS, DWE, TH, DC, TEAP, K-DV, and EP. EP, KBP, OG, and JJ contributed to the statistical analysis of the survey data. HVS conducted analysis of the RT-PCR data. EP, ASW and KBP drafted the manuscript. All authors contributed to interpretation of the study results, and revised and approved the manuscript for intellectual content. KBP and ASW are the guarantors and accept full responsibility for the work and conduct of the study, had access to the data, and controlled the decision to publish. The corresponding author (KBP) attests that all listed authors meet authorship criteria and that no others meeting the criteria have been omitted.

## Competing Interests Statement

All authors have completed the ICMJE uniform disclosure from at www.icmje.org/coi_disclore.pdf and declare: DWE declares lecture fees from Gilead, outside the submitted work; EP, PCM, NS, DWE, JIB, DC, TEAP, ASW, and KBP are employees of the University of Oxford, but not involved in the development or production of the vaccine; JIB act as an unpaid advisor to HMG on Covid but does not sit on the vaccine task force and it not involved in procurement decisions, sits on the Board of OSI who has an investment in Vaccitech who have a royalty from the ChAdOx1 vaccine when, if ever, it makes a profit; HVS reports personal fees from BioSpyder Technologies, Inc, outside the submitted work; ASW besides funding mentioned above, also received grants from Medical Research Council UK during the conduct of the study; there are no other relationships or activities that could appear to have influenced the submitted work.

## Methods

### Study participants

The Office for National Statistics (ONS) COVID-19 Infection Survey (CIS) is a large household survey with longitudinal follow-up (ISRCTN21086382, https://www.ndm.ox.ac.uk/covid-19/covid-19-infection-survey/protocol-and-information-sheets) (details in^20^). The study received ethical approval from the South Central Berkshire B Research Ethics Committee (20/SC/0195). Private households are randomly selected on a continuous basis from address lists and previous surveys to provide a representative sample across the UK. Following verbal agreement to participate, a study worker visited each selected household to take written informed consent for individuals aged 2 years and over. Parents or carers provided consent for those aged 2-15 years; those aged 10-15 years also provided written assent. All participants who completed the enrolment visit were offered a £50 voucher, and one £25 voucher for each further visit. For the current analysis we only included individuals aged 16 years and over who were potentially eligible for vaccination.

Individuals were asked about demographics, behaviours, work, and vaccination uptake (https://www.ndm.ox.ac.uk/covid-19/covid-19-infection-survey/case-record-forms). At the first visit, participants were asked for (optional) consent for follow-up visits every week for the next month, then monthly for 12 months from enrolment. At each visit, enrolled household members provided a nose and throat self-swab following instructions from the study worker, which is comparable to or even more sensitive than swabs done by healthcare workers^29^. From a random 10-20% of households, those 16 years or older were invited to provide blood monthly for antibody testing.

### Laboratory testing

Swabs were couriered directly to the UK’s national Lighthouse laboratories (Glasgow and the National Biocentre in Milton Keynes (to 8 February 2021)) where samples were tested within the national testing programme using identical methodology. The presence of three SARS-CoV-2 genes (ORF1ab, nucleocapsid protein (N), and spike protein (S)) was identified using real-time polymerase chain reaction (RT-PCR) with the TaqPath RT-PCR COVID-19 kit (Thermo Fisher Scientific, Waltham, MA, USA), analysed using UgenTec Fast Finder 3.300.5 (TagMan 2019-nCoV assay kit V2 UK NHS ABI 7500 v2.1; UgenTec, Hasselt, Belgium). The assay plugin contains an assay-specific algorithm and decision mechanism that allows conversion of the qualitative amplification assay raw data into test results with little manual intervention. Samples are called positive if either N or ORF1ab, or both, are detected. The S gene alone is not considered a reliable positive^29^, but could accompany other genes (ie, one, two, or three gene positives).

Blood samples were couriered directly to the University of Oxford, where they were tested for the SARS-CoV-2 antibody using an ELISA detecting anti-trimeric spike IgG^30^. Before 26 February 2021, the assay used fluorescence detection as previously described (positivity threshold 8 million units)^3^. After this, it used a commercialised CE-marked version of the assay, the Thermo Fisher OmniPATH 384 Combi SARS-CoV-2 IgG ELISA (Thermo Fisher Scientific, Waltham, MA, USA), with the same antigen and a colorimetric detection system (positivity threshold 42 ng/ml monoclonal antibody unit equivalents, determined from 3840 samples run in parallel).

### Inclusion and exclusion criteria

This analysis included participants aged 16 years or over (i.e. those who theoretically could have received vaccination), and all visits with positive or negative swab results from 1 December 2020 to 8 May 2021.

### Vaccination status

Participants were asked about their vaccination status at visits, including type, number of doses and date(s). Participants from England were also linked to administrative records from the National Immunisation Management Service (NIMS). We used records from NIMS where available, otherwise records from the survey, since linkage was periodic and NIMS does not contain information about vaccinations received abroad or in Northern Ireland, Scotland, and Wales. Where records were available in both, agreement on type was 98% and on dates 95% within ±7 days. The main analysis included any type of COVID vaccination, but there were only sufficient numbers to provide separate estimates for ChAdOx1 and BNT162b2 in analyses that evaluated results by vaccine type, so other vaccines were excluded from these analyses.

### SARS-CoV-2 infection episodes

PCR-positive results may be obtained at multiple visits after infection, so we grouped positive tests into ‘episodes’. Whole genome sequencing is available on only a subset of positives, and only a subsample provide monthly blood samples for antibody status, so positive episodes were defined using study PCR results. Based on the World Health Organisation (WHO) definition of re-infection as positive tests occurring at least 90 days after the onset of primary infection^31^, but also incorporating multiple consecutive negative tests, we defined the start of a new ‘infection episode’ as the date of either: i) the first PCR-positive test in the study (not preceded by any study PCR-positive test by definition); ii) a PCR-positive test after 4 or more consecutive negative tests; or iii) a PCR-positive test at least 90 days after the start of a previous infection episode with one or more negative tests immediately preceding this. Positive episodes were used to classify exposure groups and outcomes (see below).

### Exposures

At each study visit, a participant was classified into one of eight different exposure groups based on current vaccination status, study antibody and PCR tests, and (for exposure classification only) positive swab tests linked from the English national testing programme^32^ (prior to visit), as follows:

i. Visits from participants ≥21 days before first vaccination, including those currently with no vaccination date, with no prior PCR or antibody-positive in the study, nor a positive swab test in the national testing programme (as defined below) (“ Not vaccinated, not previously positive, ≥21 days before vaccination”) (baseline group);
ii. Visits from participants 1 to 21 days before first vaccination with no prior PCR or antibody-positive in the study, nor a positive swab test in the national testing programme (“ Not vaccinated, not previously positive, 1-21 days before vaccination”)
iii. Visits 0 to 7 days following a first vaccination (“ Vaccinated 0-7 days ago”);
iv. Visits 8 to 20 days following a first vaccination (“ Vaccinated 8-20 days ago”);
v. Visits 21 days or more following a first vaccination (“ ≥21 days after 1^st^ dose, no second dose”);
vi. Visits after second vaccination, ≥21 days following first vaccination (“ Post second dose”);
vii. Visits from participants that were previously PCR/antibody positive in the study, or had a positive swab test in the national testing programme and with the first positive <4 months previously and not (yet) vaccinated (“ Not vaccinated, previously positive”).
viii. Visits from participants that were previously PCR/antibody positive in the study, or had a positive swab test in the national testing programme with the first positive ≥ 4 months previously and not (yet) vaccinated.

We chose these vaccination status categories empirically based on the odds of infection episodes when modelling days since first vaccination as a continuous effect, allowing for non-linearity by using restricted cubic splines (**Extended Data Fig. 1**). Exposure group ii (Not vaccinated, not previously positive, 1-21 days before vaccination) was included because there is likely a certain degree of transient reverse causality where individuals are avoiding contact with others prior vaccination and vaccination appointments have to be rescheduled if someone test positive in the weeks before the scheduled visit (21-day cut-off reflecting the period where odds dropped below 0.50 based on **Extended Data Fig. 1**). Visits from participants that were not vaccinated, previously positive were further split by whether the first evidence of the positive test was <4 months ago or longer, because, as not everyone underwent antibody testing, by necessity our definition of a new positive episode was based on PCR positives only. More positive tests due to intermittent prolonged carriage might be expected for visits with a more recent time since the index positive episode, despite the fact that individuals were considered at risk again for a new positive episode only after having at least 4 or more consecutive negative tests or at least 90 days since the start of the index positive with at least one negative study result before the new episode. The 4 month cut-off was arbitrary being approximately the median time since the first evidence of positivity (median 125 days). As antibody status before vaccination is not available for all participants, we defined prior positivity by having either a positive antibody measurement >90 days before the visit or a previous PCR-positive episode. The choice of 90 days was arbitrary, but designed to exclude ongoing infections acquired previously being misattributed to current visits. Visits from vaccinated individuals (groups (iii)-(vi)) were defined irrespective of previous positivity (**Supplementary Table 2** & **4**) to reflect the impact of vaccination as being implemented in the UK (without regard to prior infection). Visits from the same participant were classified in different groups depending on their status at each visit.

### Outcomes

Analysis was based on visits, since these occur independently of symptoms and are therefore unbiased. Only the first test-positive visit in the first new positive infection episode starting after 1 December was used, dropping all subsequent visits in the same infection episode and all negative visits before the first time a participant could be considered “ at risk” for a new positive episode (as defined above), to avoid misattributing ongoing PCR-positivity to visit characteristics and immortal time bias respectively. Primary analysis included all first new positive infection episodes. Secondary analyses considered infection severity, by classifying positives by cycle threshold (Ct) value (<30 or ≥30) and self-reported symptoms. The threshold Ct value of 30 is somewhat arbitrary, but corresponds to ∼150 copies/ml,^26^ and is consistently used in the UK for many purposes, including algorithms for review of low level positives at the Lighthouse Laboratories where the PCR tests were performed and a threshold for attempting whole genome sequencing. For each positive test, a single Ct was calculated as the arithmetic mean across detected genes (Spearman correlation>0.98), then the minimum value was taken across positives in the infection episode to reflect the greatest measured viral burden within an episode. To allow for pre-symptomatic positives being identified in the survey, any self-reported symptoms at any visit within 0 to 35 days after the index positive in each infection episode were included (questions elicit symptoms in the last 7 days at each visit).

Finally, positive infection episodes were classified as compatible with the B.1.1.7 SARS-CoV-2 variant (those positive at least once for ORF1ab+N across the episode and never S-positive) and those that were incompatible (ORF1ab+N+S or ORF1ab+S or N+S at least once across the episode). B.1.1.7 has deletions in the S gene leading to S gene target failure, and ORF1ab+N positivity only remains a good proxy for B.1.1.7 from whole-genome sequencing from mid November 2020^33^. Positives where only a single N or single ORF1ab gene were detected were excluded from this secondary analysis.

### Confounders

The following potential confounders were adjusted for in all models as potential risk factors for acquiring SARS-CoV-2 infection: geographic area and age in years (see below), sex, ethnicity (white vs non-white as small numbers), index of multiple deprivation (percentile, calculated separately for each country in the UK)^34-37^, working in a care-home, having a patient-facing role in health or social care, presence of long-term health conditions, household size, multigenerational household, rural-urban classification^38-40^, direct or indirect contact with a hospital or care-home, smoking status, mode of travel to work, work location, and visit frequency. Details are shown in **Supplementary Table 1**.

### Statistical analysis

Associations between the different exposure groups and outcome (first positive test in an infection episode vs test-negative) were evaluated with generalised linear models with a logit link. Robust standard errors were used to account for multiple visits per-participant. To adjust for substantial confounding by calendar time and age, with non-linear effects of age which are also different by region, we included both as restricted cubic splines with knots at the 20%, 40%, 60%, and 80% percentiles of unique values and interactions between these splines and region/country (regions for England and country for Northern Ireland, Scotland and Wales). Furthermore, given previous observations of different positivity rates by age over time^20^, we added a tensor spline to model the interaction between age and calendar time with the restriction that the interaction is not doubly non-linear^41^. We considered effect modification by age of vaccination by fitting this same model, but also including an interaction between vaccine exposure group and age <75 vs ≥75 years, or long-term health conditions. There were no positives in those aged ≥75 years in one previously infected exposure group, so these groups were excluded from subgroup analysis by age. Pairwise comparisons of the exposure groups were performed using Tukey adjustments for the pairwise comparisons. Analysis was based on complete cases (>99% observations) (**Supplementary Table 2**). All statistical analyses were performed using standard functions in the following R packages available on https://cran.r-project.org: ggplot2 (version 3.3.2), rms (version 6.0-1), dplyr (version 1.0.2), emmeans (version 1.5.1), haven (version 2.3.1), sandwich (version 3.0-0), ggeffects (version 1.0.1), broom (version 0.7.2), multcomp (version 1.4-14), and Epi (version 2.44)).

## Code Availability

All statistical analyses were performed using standard functions in the following R packages: ggplot2 (version 3.3.2), rms (version 6.0-1), dplyr (version 1.0.2), emmeans (version 1.5.1), haven (version 2.3.1), sandwich (version 3.0-0), ggeffects (version 1.0.1), broom (version 0.7.2), multcomp (version 1.4-14), and Epi (version 2.44)). Code used for data analysis is available upon request.

## Notes

### Clinical Trial

ISRCTN21086382.

### Clinical Protocols

https://www.ndm.ox.ac.uk/covid-19/covid-19-infection-survey/protocol-and-information-sheets

### Author Declarations

The study received ethical approval from the South Central Berkshire B Research Ethics Committee (20/SC/0195).

