## Supplementary files for "Impact of vaccination on new SARS-CoV-2 infections in the UK"

**Supplementary Material**

**Supplementary Table 1: Characteristics of visits included in analysis**

| Characteristic [number missing] | Negative, n (%) or median (IQR) | Positive, n (%) or median (IQR) | Total, n (%) or median (IQR) |
|---|---|---|---|
| **Female [0]** | 1,034,685 (54) | 6,894 (54) | 1,041,579 (54) |
| **White ethnicity [0]** | 1,810,206 (94) | 11,577 (90) | 1,821,783 (94) |
| **Age [0]** | 55 (40, 68) | 49 (35, 61) | 55 (40, 68) |
| **Region [0]** | | | |
| London | 337,701 (17) | 3,301 (26) | 341,002 (18) |
| North_West_England | 212,234 (11) | 1,597 (12) | 213,831 (11) |
| North_East_England | 67,594 ( 3) | 473 ( 4) | 68,067 ( 3) |
| Yorkshire | 148,858 ( 8) | 828 ( 6) | 149,686 ( 8) |
| West_Midlands | 134,506 ( 7) | 868 ( 7) | 135,374 ( 7) |
| East_Midlands | 115,518 ( 6) | 752 ( 6) | 116,270 ( 6) |
| South_East_England | 237,087 (12) | 1,509 (12) | 238,596 (12) |
| South_West_England | 148,790 ( 8) | 610 ( 5) | 149,400 ( 8) |
| East_England | 184,003 (10) | 1,250 (10) | 185,253 (10) |
| Northern_Ireland | 60,847 ( 3) | 361 ( 3) | 612,08 ( 3) |
| Scotland | 181,273 ( 9) | 744 ( 6) | 182,017 ( 9) |
| Wales | 103,834 ( 5) | 533 ( 4) | 104,367 ( 5) |
| **Household size group [0]** | | | |
| One | 366,188 (19) | 2,016 (16) | 368,204 (19) |
| Two | 916,924 (47) | 5,072 (40) | 921,996 (47) |
| Three | 300,624 (16) | 2,436 (19) | 303,060 (16) |
| Four | 247,130 (13) | 2,283 (18) | 249,413 (13) |
| Five plus | 101,379 ( 5) | 1,019 ( 8) | 102,398 ( 5) |
| **Multigenerational households [0]** | 93,144 ( 5) | 814 ( 6) | 93,958 ( 5) |
| **Rural-urban classification [0]** | | | |
| Major urban | 681,144 (35) | 5,789 (45) | 686,933 (35) |
| Urban city/town | 817,866 (42) | 4954 (39) | 822,820 (42) |
| Rural town | 210,277 (11) | 1,101 ( 9) | 211,378 (11) |
| Rural village | 222,958 (12) | 982 ( 8) | 223,940 (12) |
| **Deprivation centile (1 = most deprived, 10 = least deprived) [0]** | 6 (3, 8) | 5 (3, 8) | 6 (3, 8) |
| **Ever reported to be a care home worker [0]** | 21,070 (1) | 254 ( 2) | 21,324 ( 1) |
| **Ever reported to be a patient-facing healthcare worker [0]** | 47,633 (2) | 471 ( 4) | 48,104 ( 2) |
| **Ever reported to be a person-facing social care worker [0]** | 21,855 (1) | 227 ( 2) | 22,082 ( 1) |
| **Ever reported to have a long-term health condition [0]** | 524,376 (27) | 2,961 (23) | 527,337 (27) |
| **Days since previous visit [0]** | | | |
| >14 days | 1,496,896 (77) | 9,846 (77) | 1,506,742 (77) |
| <=14 days | 348,415 (18) | 1,960 (15) | 350,375 (18) |
| Enrolment | 86,934 (4) | 1,020 ( 8) | 87,954 ( 5) |
| **Smoking status [0]** | | | |
| Non-smoker | 1,747,978 (90) | 11,655 (91) | 1,759,633 (90) |
| Tobacco smoker | 142,392 (7) | 841 ( 7) | 143,233 ( 7) |

| | | | |
|---|---|---|---|
| Only vape | 41,875 (2) | 330 ( 3) | 42,205 ( 2) |
| **Contact with hospital [7503]** | | | |
| No | 1,499,189 (78) | 9,604 (75) | 1,508,793 (78) |
| Yes, I have | 266,461 (14) | 1,988 (15) | 268,449 (14) |
| No, but someone in HH | 159,149 (8) | 1,177 ( 9) | 160,326 ( 8) |
| **Contact with care home [10814]** | | | |
| No | 1,857,072 (96) | 12,140 (95) | 1,869,212 (96) |
| Yes, I have | 38,345 (2) | 357 ( 3) | 38,702 ( 2) |
| No, but someone in HH | 26,098 (1) | 245 ( 2) | 26,343 ( 1) |
| **Work location/distancing [5759]** | | | |
| Working from home | 498,588 (26) | 3,475 (27) | 502,063 (26) |
| Working elsewhere, easy to maintain 2m | 211,157 (11) | 1,445 (11) | 212,602 (11) |
| Working elsewhere, relatively easy to maintain 2m | 98,223 (5) | 854 ( 7) | 99,077 ( 5) |
| Working elsewhere, difficult to maintain 2m | 79,833 (4) | 739 ( 6) | 80,572 ( 4) |
| Working elsewhere, very difficult to maintain 1m | 135,547 (7) | 1,390 (11) | 136,937 ( 7) |
| Employed but not currently working (including furloughed) | 75,289 (4) | 949 ( 7) | 76,238 ( 4) |
| Not employed | 124,245 (6) | 935 ( 7) | 125,180 ( 6) |
| Student | 13,029 (1) | 136 ( 1) | 13,165 ( 1) |
| Retired | 690,610 (36) | 2,868 (22) | 693,478 (36) |
| **Work travel method [5796]** | | | |
| Not travelling to work | 1,389,416 (72) | 8,260 (64) | 1,397,676 (72) |
| On foot/bike | 76,384 (4) | 611 ( 5) | 76,995 ( 4) |
| Car/taxi | 412,921 (21) | 3,435 (27) | 416,356 (21) |
| Train/bus | 46,588 (2) | 484 (4) | 47,072 ( 2) |
| Other | 1,171 (0) | 5 (0) | 1,176 ( 0) |

Note: analysis is based on visits rather than participants: factors above and vaccination exposure (detailed in Supplementary Table 2) could change over time. Numbers in square brackets in the first column indicate the number of visits with missing information.

**Supplementary Table 2: Vaccination and previous infection status for visits included in analysis**

| Vaccination and previous infection status | No prior study or national testing programme swab-positive episode and no antibody positive >90 days ago Number of visits (row %) [number of positives] | Prior study or national testing programme swab-positive episode or antibody positive >90 days ago Number of visits (row %) [number of positives] | Total visits (row %) [number of positives] |
|---|---|---|---|
| Not vaccinated, no prior positive, >21 days before vaccination | 1,012,808 (100%) [10,721] | 0 (0%) [0] | 1,012,808 (100%) [10,721] |
| Not vaccinated, no prior positive, 1-21 days before vaccination | 200,553 (100%) [643] | 0 (0%) [0] | 200,553 (100%) [643] |
| Vaccinated 0-7 days ago | 77,234 (98%) [279] | 1,929 (2%) [12] | 79,163 (100%) [291] |
| Vaccinated 8-20 days ago | 118,928 (97%) [431] | 3,443 (3%) [10] | 122,371 (100%) [441] |
| ≥21 days after 1st dose, no second dose | 355,639 (96%) [495] | 14,520 (4%) [35] | 370,159 (100%) [530] |
| Post second dose | 131,554 (96%) [83] | 6,021 (4%) [12] | 137,575 (100%) [95] |
| Not vaccinated, previously first positive <4m ago | 0 (0%) [0] | 10,534 (100%) [76] | 10,534 (100%) [76] |
| Not vaccinated, previously first positive ≥4m ago | 0 (0%) [0] | 11,908 (100%) [29] | 11,908 (100%) [29] |

Note: analysis is based on visits rather than participants: factors (Supplementary Table 1) and vaccination exposure could change over time. See methods for definition of swab-positive episodes of infection. As <4% of visits after vaccination occurred after previous infection, these were combined with those not occurring after previous infection to consider the impact of vaccination overall as implemented in the UK.

**Supplementary Table 3: Summary of key characteristics by vaccination status**

| | Not vaccinated, no prior positive, >21 days before vaccination | Not vaccinated, no prior positive, 1-21 days before vaccination | Vaccinated 0-7 days ago | Vaccinated 8-20 days ago | ≥21 days after 1st dose, no second dose | Post second dose | Not vaccinated, previously positive <4m ago | Not vaccinated, previously positive ≥4m ago | P-value* |
|---|---|---|---|---|---|---|---|---|---|
| **N (%)** | 10721 (84) | 643 (5) | 291 (2) | 441 (3) | 530 (4) | 95 (1) | 76 (1) | 29 (0) | |
| **Minimum Ct, median (IQR)** | 28.4 (20.1-32.9) | 31.5 (27.5-33.5) | 31.5 (20.8-33.8) | 30.9 (23.3-33.7) | 31.7 (26.8-33.7) | 32.7 (27.7-34.0) | 32.2 (28.8-34.1) | 33.2 (31.0-33.9) | <0.001 |
| **Gene positivity pattern** | | | | | | | | | |
| OR+N+S | 1705 (16) | 60 ( 9) | 22 ( 8) | 25 ( 6) | 25 ( 5) | 2 ( 2) | 7 ( 9) | 4 (14) | |
| OR+N (B.1.1.7 compatible) | 5628 (52) | 356 (55) | 151 (52) | 250 (57) | 314 (59) | 50 (53) | 36 (47) | 11 (38) | <0.001 |
| Other single/double | 3388 (32) | 227 (35) | 118 (41) | 166 (38) | 191 (36) | 43 (45) | 33 (43) | 14 (48) | |
| **Symptoms** | | | | | | | | | |
| None | 4492 (42) | 286 (44) | 165 (57) | 236 (54) | 323 (61) | 79 (83) | 54 (71) | 26 (90) | |
| Yes, other | 2137 (20) | 171 (27) | 53 (18) | 65 (15) | 61 (12) | 6 ( 6) | 11 (14) | 2 ( 7) | <0.001 |
| Yes, cough/fever | 4083 (38) | 184 (29) | 71 (24) | 138 (31) | 145 (27) | 10 (11) | 10 (13) | 1 ( 3) | |
| **Ct/ symptoms combination** | | | | | | | | | |
| Ct <30 and symptoms reported | 4235 (40) | 163 (25) | 77 (26) | 134 (30) | 105 (20) | 8 ( 8) | 9 (12) | 0 ( 0) | |
| Ct <30 and no symptoms reported | 1788 (17) | 84 (13) | 51 (18) | 68 (15) | 96 (18) | 19 (20) | 14 (18) | 5 (17) | |
| Ct 30+ and symptoms reported | 1985 (19) | 192 (30) | 47 (16) | 69 (16) | 101 (19) | 8 ( 8) | 12 (16) | 3 (10) | |
| Ct 30+ and no symptoms reported | 2713 (25) | 204 (32) | 116 (40) | 170 (39) | 228 (43) | 60 (63) | 41 (54) | 21 (72) | |
| Visit with prior negative result | | | 267 (92) | 409 (93) | 494 (93) | 87 (92) | | | |

*p-values calculated using two-sided Kruskal-Wallis test for continuous factors and chi-squared test for categorical factors and indicate differences between each vaccination status group

Note: showing n (col %) or median IQR

**Supplementary Table 4: Odds ratios (OR) from main models**

| Vaccination status | | Outcome: All positives | | Outcome: split by Ct | | Outcome: split by symptoms | | Outcome: Split by gene positivity pattern | |
|---|---|---|---|---|---|---|---|---|---|
| | | Unadjusted | Adjusted | Mean Ct <30 | Mean Ct ≥30 | Symptoms reported | No symptoms reported | S gene positive (plus 1 or 2 other genes) | B.1.1.7 compatible (ORFab+N) |
| **Not vaccinated, no prior positive, 1-21 days before vaccination** | OR (95% CI) | 0.30 (0.28, 0.33) | 0.28 (0.25, 0.30) | 0.21 (0.18, 0.24) | 0.35 (0.31, 0.40) | 0.29 (0.26, 0.32) | 0.27 (0.24, 0.31) | 0.25 (0.19, 0.33) | 0.28 (0.25, 0.32) |
| | P-value vs baseline | <0.001 | <0.001 | <0.001 | <0.001 | <0.001 | <0.001 | <0.001 | <0.001 |
| **Vaccinated 0-7 days ago** | OR (95% CI) | 0.34 (0.31, 0.39) | 0.37 (0.32, 0.42) | 0.32 (0.26, 0.39) | 0.42 (0.36, 0.51) | 0.29 (0.24, 0.35) | 0.48 (0.40, 0.57) | 0.31 (0.20, 0.48) | 0.34 (0.28, 0.40) |
| | P-value vs baseline | <0.001 | <0.001 | <0.001 | <0.001 | <0.001 | <0.001 | <0.001 | <0.001 |
| | Pairwise p-value vs group above | 0.463 | 0.002 | 0.002 | 0.460 | 1.00 | <0.001 | 0.993 | 0.580 |
| **Vaccinated 8-20 days ago** | OR (95% CI) | 0.34 (0.31, 0.37) | 0.44 (0.39, 0.49) | 0.39 (0.33, 0.47) | 0.50 (0.42, 0.58) | 0.37 (0.31, 0.43) | 0.55 (0.46, 0.65) | 0.33 (0.21, 0.51) | 0.42 (0.36, 0.49) |
| | P-value vs baseline | <0.001 | <0.001 | <0.001 | <0.001 | <0.001 | <0.001 | <0.001 | <0.001 |
| | Pairwise p-value vs group above | 1.00 | 0.251 | 0.524 | 0.767 | 0.396 | 0.856 | 1.00 | 0.347 |
| **≥21 days after 1st dose, no second dose** | OR (95% CI) | 0.13 (0.12, 0.15) | 0.36 (0.32, 0.41) | 0.25 (0.21, 0.31) | 0.50 (0.42, 0.59) | 0.24 (0.20, 0.30) | 0.54 (0.45, 0.64) | 0.39 (0.24, 0.63) | 0.32 (0.27, 0.38) |
| | P-value vs baseline | <0.001 | <0.001 | <0.001 | <0.001 | <0.001 | <0.001 | <0.001 | <0.001 |
| | Pairwise p-value vs group above | <0.001 | 0.065 | 0.001 | 1.00 | 0.002 | 1.00 | 0.997 | 0.063 |
| **Post second dose** | OR (95% CI) | 0.06 (0.05, 0.08) | 0.20 (0.16, 0.26) | 0.09 (0.06, 0.15) | 0.36 (0.26, 0.50) | 0.05 (0.03, 0.10) | 0.42 (0.31, 0.57) | 0.06 (0.01, 0.32) | 0.15 (0.11, 0.21) |
| | P-value vs baseline | <0.001 | <0.001 | <0.001 | <0.001 | <0.001 | <0.001 | 0.001 | <0.001 |
| | Pairwise p-value vs group above | <0.001 | <0.001 | <0.001 | 0.376 | <0.001 | <0.001 | <0.001 | <0.001 |
| | OR (95% CI) | 0.68 (0.54, 0.85) | 0.72 (0.58, 0.91) | 0.39 (0.26, 0.58) | 1.17 (0.89, 1.54) | 0.33 (0.22, 0.51) | 1.34 (1.02, 1.76) | 0.53 (0.25, 1.13) | 0.58 (0.42, 0.81) |

| Vaccination status | | Outcome: All positives | | Outcome: split by Ct | | Outcome: split by symptoms | | Outcome: Split by gene positivity pattern | |
|---|---|---|---|---|---|---|---|---|---|
| | | Unadjusted | Adjusted | Mean Ct <30 | Mean Ct ≥30 | Symptoms reported | No symptoms reported | S gene positive (plus 1 or 2 other genes) | B.1.1.7 compatible (ORFab+N) |
| **Not vaccinated, previously positive <4m ago** | P-value vs baseline | 0.001 | 0.006 | <0.001 | 0.272 | <0.001 | 0.035 | 0.099 | 0.001 |
| | Pairwise p-value vs group above | <0.001 | <0.001 | <0.001 | <0.001 | <0.001 | <0.001 | 0.214 | <0.001 |
| **Not vaccinated, previously positive, ≥4m ago** | OR (95% CI) | 0.23 (0.16, 0.33) | 0.31 (0.21, 0.45) | 0.09 (0.04, 0.21) | 0.63 (0.42, 0.95) | 0.05 (0.02, 0.16) | 0.72 (0.49, 1.07) | 0.48 (0.22, 1.08) | 0.19 (0.10, 0.34) |
| | P-value vs baseline | <0.001 | <0.001 | <0.001 | 0.027 | <0.001 | 0.104 | 0.075 | <0.001 |
| | Pairwise p-value vs second dose | <0.001 | 0.523 | 1.00 | 0.330 | 1.00 | 0.338 | 0.316 | 0.997 |

Note: all odds ratios are compared to the baseline (reference) group of **Not vaccinated, no prior positive, >21 days before vaccination.** Results shown graphically in **Figure 2**. P-values vs baseline use a two-sided Wald test vs the baseline group. Pairwise p-values are two-sided and are a Wald test of whether the OR for each vaccine status group is different to the vaccine status group immediately above; so respectively "Vaccinated 0 to 7 days ago, 1 dose" vs "Not vaccinated, no prior positive, 1-21 days before vaccination", "Vaccinated 8 to 20 days ago" vs "Vaccinated 0 to 7 days ago", "Vaccinated ≥ 21 days ago, 2 doses" vs "Vaccinated ≥ 21 days ago, 1 dose" and "Not vaccinated, but positive < or >4m ago" vs "Vaccinated ≥ 21 days ago, 2 doses".

**Supplementary Table 5: Odds ratios of other factors in main model for new PCR-positives**

| Term | Odds ratio | 95% CI | | P-value |
|---|---|---|---|---|
| Contact care home | | | | |
| No | 1 | | | |
| No, but someone in HH | 1.33 | 1.17 | 1.51 | <0.001 |
| Yes, I have | 1.33 | 1.18 | 1.49 | <0.001 |
| Contact hospital | | | | |
| No | 1 | | | |
| No, but someone in HH | 1.11 | 1.04 | 1.18 | 0.075 |
| Yes, I have | 1.30 | 1.24 | 1.37 | <0.001 |
| Vaccination status | | | | |
| Not vaccinated, no prior positive, >21 days before vaccination | 1 | | | |
| Not vaccinated, no prior positive, 1-21 days before vaccination | 0.28 | 0.25 | 0.30 | <0.001 |
| Vaccinated 0 to 7 days ago | 0.37 | 0.32 | 0.42 | <0.001 |
| Vaccinated 8 to 20 days ago | 0.44 | 0.39 | 0.49 | <0.001 |
| ≥21 days since 1st vaccination, no second dose | 0.36 | 0.32 | 0.41 | <0.001 |
| Post second dose | 0.20 | 0.16 | 0.26 | <0.001 |
| Not vaccinated, prior positive <4 months ago | 0.72 | 0.58 | 0.91 | 0.006 |
| Not vaccinated, prior positive ≥4 months ago | 0.31 | 0.21 | 0.45 | <0.001 |
| Ethnicity | | | | |
| White | 1 | | | |
| Non-White | 1.13 | 1.06 | 1.20 | <0.001 |
| Ever care home worker | | | | |
| No | 1 | | | |
| Yes | 1.34 | 1.16 | 1.55 | <0.001 |
| Ever reported long-term health conditions | | | | |
| No | 1 | | | |
| Yes | 0.99 | 0.94 | 1.03 | 0.524 |
| Ever patient facing healthcare worker | | | | |
| No | 1 | | | |
| yes | 1.51 | 1.36 | 1.67 | <0.001 |
| Ever person-facing social care worker | | | | |
| No | 1 | | | |
| Yes | 1.46 | 1.27 | 1.69 | <0.001 |
| Household size | | | | |
| One | 1 | | | |
| Two | 1.00 | 0.95 | 1.05 | 0.994 |
| Three | 1.20 | 1.12 | 1.28 | <0.001 |
| Four | 1.34 | 1.25 | 1.43 | <0.001 |
| Five_plus | 1.43 | 1.32 | 1.56 | <0.001 |
| Deprivation score | 0.96 | 0.95 | 0.97 | <0.001 |
| Multigenerational household | | | | |
| No | 1 | | | |

| Term | Odds ratio | 95% CI | | P-value |
|---|---|---|---|---|
| Yes | 0.96 | 0.89 | 1.04 | 0.314 |
| Sex | | | | |
| Male | 1 | | | |
| Female | 0.99 | 0.96 | 1.03 | 0.670 |
| Smoking status | | | | |
| Non-smoker | 1 | | | |
| Only vape | 1.03 | 0.92 | 1.15 | 0.641 |
| Tobacco smoker | 0.79 | 0.73 | 0.85 | <0.001 |
| Days since last visit | | | | |
| >14 days since last visit | 1 | | | |
| <=14 days since last visit | 0.56 | 0.53 | 0.59 | <0.001 |
| Enrolment | 1.01 | 0.94 | 1.08 | 0.754 |
| Work location distancing | | | | |
| Working from home | 1 | | | |
| Working elsewhere, difficult to maintain 2m | 1.32 | 1.02 | 1.71 | 0.037 |
| Working elsewhere, easy to maintain 2m | 1.00 | 0.78 | 1.29 | 0.979 |
| Working elsewhere, relatively easy to maintain 2m | 1.26 | 0.97 | 1.63 | 0.083 |
| Working elsewhere, very difficult to maintain 1m | 1.51 | 1.17 | 1.94 | 0.002 |
| Furloughed/temporarily not working | 1.82 | 1.69 | 1.96 | <0.001 |
| Retired | 1.00 | 0.93 | 1.08 | 0.997 |
| Student | 1.04 | 0.82 | 1.31 | 0.745 |
| Not working | 1.05 | 0.97 | 1.13 | 0.249 |
| Work travel method | | | | |
| Not travelling to work | 1 | | | |
| Car/taxi | 1.20 | 0.94 | 1.54 | 0.148 |
| On foot/bike | 1.06 | 0.82 | 1.37 | 0.646 |
| Other | 0.51 | 0.21 | 1.27 | 0.151 |
| Train/bus | 1.19 | 0.92 | 1.54 | 0.194 |
| Rural urban classification | | | | |
| Major urban area | 1 | | | |
| Urban city/town | 0.85 | 0.81 | 0.90 | <0.001 |
| Rural town | 0.79 | 0.73 | 0.85 | <0.001 |
| Rural village | 0.67 | 0.62 | 0.73 | <0.001 |

[1]Interactions included in model: study day by age, study day by region, age by region.

Note: analysis is based on visits rather than participants: factors and vaccination exposure could change over time. Two-sided p-values from Wald test.

**Supplementary Table 6: Odds ratios from models split by vaccine type**

| Vaccination status | | Outcome: All positives | Outcome: split by Ct | | Outcome: split by symptoms | |
|---|---|---|---|---|---|---|
| | | Adjusted | Mean Ct <30 | Mean Ct ≥30 | Symptoms reported | No symptoms reported |
| **Not vaccinated, no prior positive, 1-21 days before vaccination** | OR (95% CI) | 0.28 (0.25, 0.30) | 0.21 (0.18, 0.24) | 0.35 (0.31, 0.40) | 0.29 (0.26, 0.32) | 0.27 (0.24, 0.31) |
| | P-value vs baseline | <0.001 | <0.001 | <0.001 | <0.001 | <0.001 |
| **Vaccinated with ChAdOx1 0-7 days ago** | OR (95% CI) | 0.33 (0.27, 0.40) | 0.24 (0.17, 0.32) | 0.43 (0.34, 0.54) | 0.24 (0.18, 0.32) | 0.45 (0.35, 0.58) |
| | P-value vs baseline | <0.001 | <0.001 | <0.001 | <0.001 | <0.001 |
| | Pairwise p-value vs group above | 0.799 | 1.00 | 0.907 | 0.986 | 0.006 |
| **Vaccinated with BNT162b2 0-7 days ago** | OR (95% CI) | 0.40 (0.34, 0.47) | 0.38 (0.30, 0.48) | 0.43 (0.34, 0.54) | 0.33 (0.26, 0.42) | 0.51 (0.40, 0.63) |
| | P-value vs baseline | <0.001 | <0.001 | <0.001 | <0.001 | <0.001 |
| | Pairwise p-value vs group above | 0.882 | 0.269 | 1.00 | 0.834 | 1.00 |
| **Vaccinated with ChAdOx1 8-20 days ago** | OR (95% CI) | 0.43 (0.36, 0.50) | 0.37 (0.29, 0.48) | 0.49 (0.39, 0.61) | 0.38 (0.30, 0.49) | 0.50 (0.40, 0.64) |
| | P-value vs baseline | <0.001 | <0.001 | <0.001 | <0.001 | <0.001 |
| | Pairwise p-value vs group above | 1.00 | 1.00 | 0.999 | 0.998 | 1.00 |
| **Vaccinated with BNT162b2 8-20 days ago** | OR (95% CI) | 0.45 (0.39, 0.51) | 0.40 (0.33, 0.49) | 0.51 (0.42, 0.61) | 0.36 (0.29, 0.44) | 0.58 (0.48, 0.70) |
| | P-value vs baseline | <0.001 | <0.001 | <0.001 | <0.001 | <0.001 |
| | Pairwise p-value vs group above | 1.00 | 1.00 | 1.00 | 1.00 | 0.994 |
| **≥21 days after 1st ChAdOx1 dose, no second dose** | OR (95% CI) | 0.39 (0.32, 0.46) | 0.27 (0.20, 0.35) | 0.54 (0.42, 0.68) | 0.29 (0.22, 0.38) | 0.53 (0.42, 0.67) |
| | P-value vs baseline | <0.001 | <0.001 | <0.001 | <0.001 | <0.001 |
| | Pairwise p-value vs group above | 0.94 | 0.255 | 1.00 | 0.976 | 1.00 |
| **≥21 days after 1st BNT162b2 dose, no second dose** | OR (95% CI) | 0.34 (0.29, 0.40) | 0.24 (0.19, 0.30) | 0.47 (0.39, 0.58) | 0.22 (0.17, 0.28) | 0.53 (0.43, 0.65) |
| | P-value vs baseline | <0.001 | <0.001 | <0.001 | <0.001 | <0.001 |
| | Pairwise p-value vs group above | 0.97 | 1.00 | 0.995 | 0.724 | 1.00 |
| **Post second ChAdOx1 dose** | OR (95% CI) | 0.21 (0.12, 0.35) | 0.12 (0.05, 0.28) | 0.33 (0.17, 0.65) | 0.08 (0.03, 0.22) | 0.39 (0.21, 0.73) |
| | P-value vs baseline | <0.001 | <0.001 | 0.001 | <0.001 | 0.003 |
| | Pairwise p-value vs group above | 0.709 | 0.867 | 0.996 | 0.637 | 0.998 |
| **Post second BNT162b2 dose** | OR (95% CI) | 0.20 (0.15, 0.26) | 0.08 (0.05, 0.14) | 0.37 (0.26, 0.51) | 0.05 (0.02, 0.09) | 0.42 (0.31, 0.57) |
| | P-value vs baseline | <0.001 | <0.001 | <0.001 | <0.001 | <0.001 |
| | Pairwise p-value vs group above | 1.00 | 0.999 | 1.00 | 0.996 | 1.00 |
| **Not vaccinated, prior positive <4m ago** | OR (95% CI) | 0.72 (0.57, 0.91) | 0.39 (0.26, 0.58) | 1.17 (0.89, 1.54) | 0.33 (0.22, 0.51) | 1.34 (1.02, 1.75) |
| | P-value vs baseline | 0.006 | <0.001 | 0.271 | <0.001 | 0.036 |
| | Pairwise p-value vs group above | <0.001 | <0.001 | <0.001 | <0.001 | <0.001 |

| Vaccination status | | Outcome: All positives | Outcome: split by Ct | | Outcome: split by symptoms | |
|---|---|---|---|---|---|---|
| | | Adjusted | Mean Ct <30 | Mean Ct ≥30 | Symptoms reported | No symptoms reported |
| **Not vaccinated, prior positive ≥4m ago** | OR (95% CI) | 0.31 (0.21, 0.45) | 0.09 (0.04, 0.21) | 0.63 (0.42, 0.95) | 0.05 (0.02, 0.16) | 0.72 (0.49, 1.07) |
| | P-value vs baseline | <0.001 | <0.001 | 0.027 | <0.001 | 0.102 |
| | Pairwise p-value vs post second BNT162b2 dose | 0.704 | 1.00 | 0.570 | 1.00 | 0.531 |

Note: all odds ratios are compared to the base category of Not vaccinated, no prior positive, >21 days before vaccination. Results shown graphically in **Figure 4**. P-values vs baseline use a two-sided Wald test vs the baseline group. Pairwise p-values are two-sided and are a Wald test of whether the specified OR vaccine status is different to the vaccine status group above

**Supplementary Table 7: Sensitivity analysis for all new PCR-positives excluding potential confounders that are may also be effected by vaccination or prior infection status**

| Vaccination status | Main model (all covariates)<br><br>OR (95% CI) | Model without 'Work location/ distancing' and 'work travel method'<br><br>OR (95% CI) | Model without 'Work location/ distancing' and 'work travel method' and 'Contact care home'<br>OR (95% CI) |
|---|---|---|---|
| **Not vaccinated, no prior positive, 1-21 days before vaccination** | 0.28 (0.25, 0.30) | 0.28 (0.26, 0.31) | 0.29 (0.26, 0.31) |
| **Vaccinated 0-7 days ago** | 0.37 (0.32, 0.42) | 0.38 (0.34, 0.44) | 0.39 (0.34, 0.44) |
| **Vaccinated 8-20 days ago** | 0.44 (0.39, 0.49) | 0.47 (0.41, 0.52) | 0.47 (0.42, 0.53) |
| **≥21 days after 1st dose, no second dose** | 0.36 (0.32, 0.41) | 0.39 (0.34, 0.45) | 0.40 (0.35, 0.46) |
| **Post second dose** | 0.20 (0.16, 0.26) | 0.23 (0.18, 0.30) | 0.23 (0.18, 0.30) |
| **Not vaccinated, prior positive <4m ago** | 0.72 (0.58, 0.91) | 0.74 (0.59, 0.93) | 0.74 (0.59, 0.93) |
| **Not vaccinated, prior positive ≥4m ago** | 0.31 (0.21, 0.45) | 0.32 (0.22, 0.46) | 0.32 (0.22, 0.46) |

**Supplementary Table 8: Number of complete cases included in main models**

| **Model** | **Positive swabs, n (%)** | **Negative swabs, n (%)** | **Total, n (%)** |
|---|---|---|---|
| **Outcome: All positives** | 12,705 (99) | 1,915,184 (99) | 1,927,889 (99) |
| **Outcome: Positives based on Ct values** | | | |
| Ct <30 | 6,801 (99) | 1,921,088 (99) | 1,927,889 (99) |
| Ct 30+ | 5,904 (99) | 1,921,985 (99) | 1,927,889 (99) |
| **Outcome: Positives based on symptoms** | | | |
| Symptoms reported | 7,088 (99) | 1,920,801 (99) | 1,927,889 (99) |
| No symptoms reported | 5,617 (99) | 1,922,272 (99) | 1,927,889 (99) |
| **Outcome: Positives based on Ct pattern** | | | |
| S gene positive (plus 1 or 2 other genes) | 1,941 (99) | 1,925,948 (99) | 1,927,889 (99) |
| B.1.1.7 compatible (ORFab+N) | 6,734 (99) | 1,921,155 (99) | 1,927,889 (99) |

**Supplementary Table 9: Number of participants and visits included in models split by vaccine type**

| Vaccination status | Number of participants, n (%*) | Positive visits, n (%) | Negative visits, n (%) | Total, n (%) |
|---|---|---|---|---|
| **Total** | 383,812 (100) | 12,826 (100) | 1,932,245 (100) | 1,945,071 (100) |
| **Not vaccinated, no prior positive, >21 days before vaccination** | 329,419 (86) | 10,721 (84) | 1,002,087 (52) | 1,012,808 (52) |
| **Not vaccinated, no prior positive, 1-21 days before vaccination** | 176,296 (46) | 643 ( 5) | 199,910 (10) | 200,553 (10) |
| **Vaccinated with ChAdOx1 0-7 days** | 50,010 (13) | 121 ( 1) | 50,717 ( 3) | 50,838 ( 3) |
| **Vaccinated with BNT162b2 0-7 days** | 26,734 ( 7) | 168 ( 1) | 27,224 ( 1) | 27,392 ( 1) |
| **Vaccinated with ChAdOx1 8-20 days** | 73,840 (19) | 177 ( 1) | 76,872 ( 4) | 77,049 ( 4) |
| **Vaccinated with BNT162b2 8-20 days** | 41,004 (11) | 261 ( 2) | 43,934 ( 2) | 44,195 ( 2) |
| **≥21 days after 1$^{st}$ ChAdOx1 dose, no second dose** | 144,859 (38) | 236 ( 2) | 222,400 (12) | 222,636 (11) |
| **≥21 days after 1$^{st}$ BNT162b2 dose, no second dose** | 81,171 (21) | 289 ( 2) | 144,947 ( 8) | 145,236 (7) |
| **Post second ChAdOx1 dose** | 41,018 (11) | 21 ( 0) | 47,392 ( 2) | 47,413 ( 2) |
| **Post second BNT162b2 dose** | 57,646 (15) | 72 ( 1) | 89,185 ( 5) | 89,257 ( 5) |
| **Not vaccinated, prior positive <4m ago** | 8,849 ( 2) | 76 ( 1) | 10,458 ( 1) | 10,534 ( 1) |
| **Not vaccinated, prior positive ≥4m ago** | 6,468 ( 2) | 29 ( 0) | 11,879 ( 1) | 11,908 ( 1) |
| **Other vaccinated administered (not included in model)** | 2,173 ( 1) | 12 ( 0) | 5,240 ( 0) | 5,252 ( 0) |

*Percentage of participants does not add to 100 because each participant has multiple visits which can occur in multiple vaccination status categories. For participants, percentages show the percentage of participants contributing any visit to each category.

**Supplementary Table 10: Number of participants and visits included in subgroup analyses of all new PCR-positives ny age (A) and long term health conditions (B)**

### (A) Age

| Vaccination status | Age <75 | | | | Age 75+ | | | |
|---|---|---|---|---|---|---|---|---|
| | Number of participants, n (%*) | Positive visits, n (%) | Negative visits, n (%) | Total visits | Number of participants, n (%*) | Positive visits, n (%) | Negative visits, n (%) | Total visits |
| **Not vaccinated, no prior positive, >21 days before vaccination** | 303874 (79) | 10338 (1) | 962432 (99) | 972770 (100) | 25665 ( 7) | 383 ( 1) | 39655 (99) | 40038 (100) |
| **Not vaccinated, no prior positive, 1-21 days before vaccination** | 151900 (40) | 536 (0) | 171516 (100) | 172052 (100) | 24411 ( 6) | 107 ( 0) | 28394 (100) | 28501 (100) |
| **Vaccinated 0-7 days ago** | 66450 (17) | 231 (0) | 67400 (100) | 67631 (100) | 11194 ( 3) | 60 ( 1) | 11472 (99) | 11532 (100) |
| **Vaccinated 8-20 days ago** | 98597 (26) | 340 (0) | 103141 (100) | 103481 (100) | 17270 ( 4) | 101 ( 1) | 18789 (99) | 18890 (100) |
| **≥21 days after 1st dose, no second dose** | 193680 (50) | 424 (0) | 304777 (100) | 305201 (100) | 33937 ( 9) | 106 ( 0) | 64852 (100) | 64958 (100) |
| **Post second dose** | 67700 (18) | 50 (0) | 88653 (100) | 88703 (100) | 31622 ( 8) | 45 (0) | 48827 (100) | 48872 (100) |

### (B) Long-term health conditions

| Vaccination status | Never reported long-term health conditions | | | | Ever reported long-term health conditions | | | |
|---|---|---|---|---|---|---|---|---|
| | Number of participants, n (%*) | Positive swabs, n (%) | Negative swabs, n (%) | Total | Number of participants, n (%*) | Positive swabs, n (%) | Negative swabs, n (%) | Total |
| **Not vaccinated, no prior positive, >21 days before vaccination** | 244816 (64) | 8443 ( 1) | 786961 (99) | 795404 (100) | 84603 (22) | 2278 ( 1) | 215126 (99) | 217404 (100) |
| **Not vaccinated, no prior positive, 1-21 days before vaccination** | 120409 (31) | 418 ( 0) | 136227 (100) | 136645 (100) | 55887 (15) | 225 ( 0) | 63683 (100) | 63908 (100) |
| **Vaccinated 0-7 days ago** | 53328 (14) | 201 ( 0) | 54117 (100) | 54318 (100) | 24314 ( 6) | 90 ( 0) | 24755 (100) | 24845 (100) |
| **Vaccinated 8-20 days ago** | 78691 (21) | 296 ( 0) | 82751 (100) | 83047 (100) | 37175 (10) | 145 ( 0) | 39179 (100) | 39324 (100) |
| **≥21 days after 1st dose, no second dose** | 152346 (40) | 359 ( 0) | 243036 (100) | 243395 (100) | 74906 (20) | 171 ( 0) | 126593 (100) | 126764 (100) |
| **Post second dose** | 62191 (16) | 61 ( 0) | 86155 (100) | 86216 (100) | 37076 (10) | 34 ( 0) | 51325 (100) | 51359 (100) |
| **Not vaccinated, prior positive <4m ago** | 7345 ( 2) | 62 ( 1) | 8684 (99) | 8746 (100) | 1504 ( 0) | 14 ( 1) | 1774 (99) | 1788 (100) |
| **Not vaccinated, prior positive ≥4m ago** | 5421 ( 1) | 25 ( 0) | 9938 (100) | 9963 (100) | 1047 ( 0) | 4 ( 0) | 1941 (100) | 1945 (100) |

* Percentage of participants does not add to 100 because each participant has multiple visits which can occur in multiple vaccination status categories. For participants, percentages show the percentage of participants contributing any visit to each category.
